# Effectiveness of BNT162b2 COVID-19 Vaccination in Children Aged 5–17 Years in the United States

**DOI:** 10.1101/2023.09.06.23294426

**Authors:** Rachel P. Ogilvie, J. Bradley Layton, Patricia C. Lloyd, Yixin Jiao, Djeneba Audrey Djibo, Hui Lee Wong, Joann F. Gruber, Ron Parambi, Jie Deng, Michael Miller, Jennifer Song, Lisa B. Weatherby, Lauren Peetluk, An-Chi Lo, Kathryn Matuska, Michael Wernecke, Christine L. Bui, Tainya C. Clarke, Sylvia Cho, Elizabeth J. Bell, Grace Yang, Kandace L. Amend, Richard A. Forshee, Steven A. Anderson, Cheryl N McMahill-Walraven, Yoganand Chillarige, Mary S. Anthony, John D. Seeger, Azadeh Shoaibi

**Author notes:** Co-first authors. **Corresponding Author:** J. Bradley Layton 3040 East Cornwallis Rd, PO Box 12194 Research Triangle Park, NC 27709.

## Abstract

**Importance:** COVID-19 vaccines are authorized for use in children in the United States; real-world assessment of vaccine effectiveness in children is needed.

**Objective:** To estimate the effectiveness of receiving a complete primary series of monovalent BNT162b2 (Pfizer-BioNTech) COVID-19 vaccine in US children.

**Design:** A cohort study of children aged 5–17 years vaccinated with BNT162b2 matched with unvaccinated children.

**Setting:** Participants identified in Optum and CVS Health insurance administrative claims databases were linked with Immunization Information System (IIS) COVID-19 vaccination records from 16 US jurisdictions between December 11, 2020, and May 31, 2022 (end date varied by database and IIS).

**Participants:** Vaccinated children were followed from their first BNT162b2 dose and matched to unvaccinated children on calendar date, US county of residence, and demographic and clinical factors. Censoring occurred if vaccinated children failed to receive a timely dose 2 or if unvaccinated children received any dose.

**Exposure:** BNT162b2 vaccinations were identified using IIS vaccination records and insurance claims.

**Main Outcomes and Measures:** Two COVID-19 outcome definitions were evaluated: COVID-19 diagnosis in any medical setting and COVID-19 diagnosis in hospitals/emergency departments (EDs). Propensity score-weighted hazard ratios (HRs) and 95% confidence intervals (CIs) were estimated with Cox proportional hazards models, and vaccine effectiveness (VE) was estimated as 1 minus HR. VE was estimated overall, within age subgroups, and within variant-specific eras. Sensitivity, negative control, and quantitative bias analyses evaluated various potential biases.

**Results:** There were 453,655 eligible vaccinated children one-to-one matched to unvaccinated comparators (mean age 12 years; 50% female). COVID-19 hospitalizations/ED visits were rare in children, regardless of vaccination status (Optum, 41.2 per 10,000 person-years; CVS Health, 44.1 per 10,000 person-years). Overall, vaccination was associated with reduced incidence of any medically diagnosed COVID-19 (meta-analyzed VE = 38% [95% CI, 36%-40%]) and hospital/ED–diagnosed COVID-19 (meta-analyzed VE = 61% [95% CI, 56%-65%]). VE estimates were lowest among children 5–11 years and during the omicron variant era.

**Conclusions and Relevance:** Receipt of a complete BNT162b2 vaccine primary series was associated with overall reduced medically diagnosed COVID-19 and hospital/ED–diagnosed COVID-19 in children; observed VE estimates differed by age group and variant era.

**KEY POINTS:** *Question:* Does receiving a complete primary series of monovalent BNT162b2 COVID-19 vaccine reduce COVID-19 diagnoses and ED visits/hospitalizations in children aged 5–17 years?

*Findings:* In this cohort study evaluating vaccination records and medical encounters from 827,149 children, recipients of a complete primary series of BNT162b2 generally had lower rates of COVID-19 diagnoses and ED visits/hospitalizations than unvaccinated children. Vaccine effectiveness was lower in children aged 5–11 years and during omicron variant predominance.

*Meaning:* Receiving a primary series of monovalent BNT162b2 COVID-19 vaccine is effective in preventing COVID-19 diagnoses; changing variants and younger age groups may require further evaluations.

## INTRODUCTION

The burden of coronavirus disease 2019 (COVID-19) has been relatively mild among children,^1^ but COVID-19 hospitalizations increased among children as new SARS-CoV-2 variants circulated.^2^ At the time of study execution, monovalent BNT162b2 (Pfizer-BioNTech’s messenger ribonucleic acid [mRNA] COVID-19 vaccine, Comirnaty^®^) was authorized for children aged 5–17 years. In preauthorization trials, BNT162b2 demonstrated immunogenicity and efficacy in preventing confirmed COVID-19 infection in children and adolescents.^3,4^ However, compared with adults, there are fewer available data on the real-world effectiveness of COVID-19 vaccination in US children.

The effectiveness of COVID-19 vaccines has been evaluated in different US geographic and healthcare settings, time periods, and age groups using an array of study designs and data sources.^5–10^ As part of its continued surveillance of authorized vaccines, the US Food and Drug Administration (FDA) Biologics Effectiveness and Safety (BEST) Initiative evaluated the real-world effectiveness of monovalent BNT162b2 in US children using national insurance claims databases linked to immunization information system (IIS) vaccination records to improve vaccine capture and limit bias. The primary objective was to assess the effectiveness of receiving a complete primary series of monovalent BNT162b2 COVID-19 vaccination compared with being unvaccinated in preventing medically diagnosed COVID-19 and hospital/emergency department (ED)–diagnosed COVID-19 in children aged 5–17 years. Secondary objectives assessed vaccine effectiveness by age subgroup and variant era.

## MATERIALS AND METHODS

### Population and Data Source

This cohort study used two commercial insurance administrative claims data sources: Optum pre-adjudicated claims and CVS Health adjudicated claims databases (Supplemental Methods: Data Sources). To enhance vaccine administration capture, claims databases were supplemented with IIS COVID-19 vaccination records^11,12^ (Optum, 10 IIS from 10 US states; CVS Health, 11 IIS from 9 US states). Each data source’s study population was restricted to geographic areas of overlap between claims and IIS data.

The study period started on December 11, 2020, when BNT162b2 was authorized for ages 16–17 years, and ended at the latest complete IIS data for each jurisdiction. In Optum, end dates varied by IIS, ranging from September 30, 2021, to May 31, 2022; in CVS Health, the end date was March 31, 2022 for all IISs. BNT162b2 was the only COVID-19 vaccine authorized for children aged 5–17 years during the study period.

Vaccinated children were identified at their first recorded COVID-19 vaccine dose during the study period; children with non-BNT162b2, brand-unspecified, or unclassifiable COVID-19 vaccine records as the first observed dose were excluded. The date of dose 1 was assigned as Time 0 in vaccinated children (eFigure 1). Unvaccinated children were one-to-one exact matched with replacement on calendar date and the following: age groups (5–11, 12–15, 16–17 years), sex, county of residence, immunocompromised status, pregnancy status, previous COVID-19 diagnosis, having a comorbidity increasing the risk of severe COVID-19,^13^ and influenza vaccine receipt in the previous year. The calendar date of dose 1 for the vaccinated child was set as Time 0 for the matched unvaccinated child.

**Figure 1.**
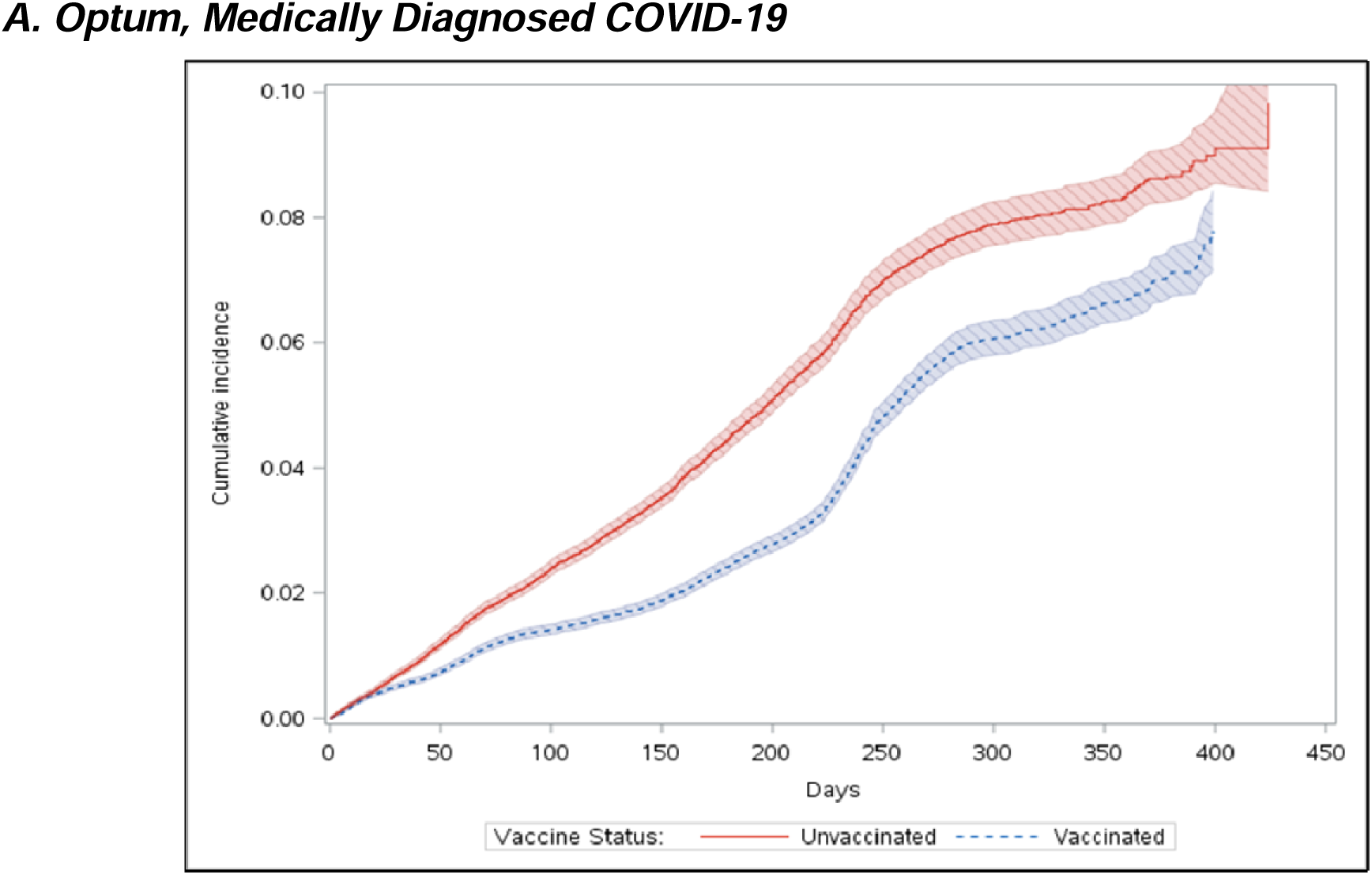
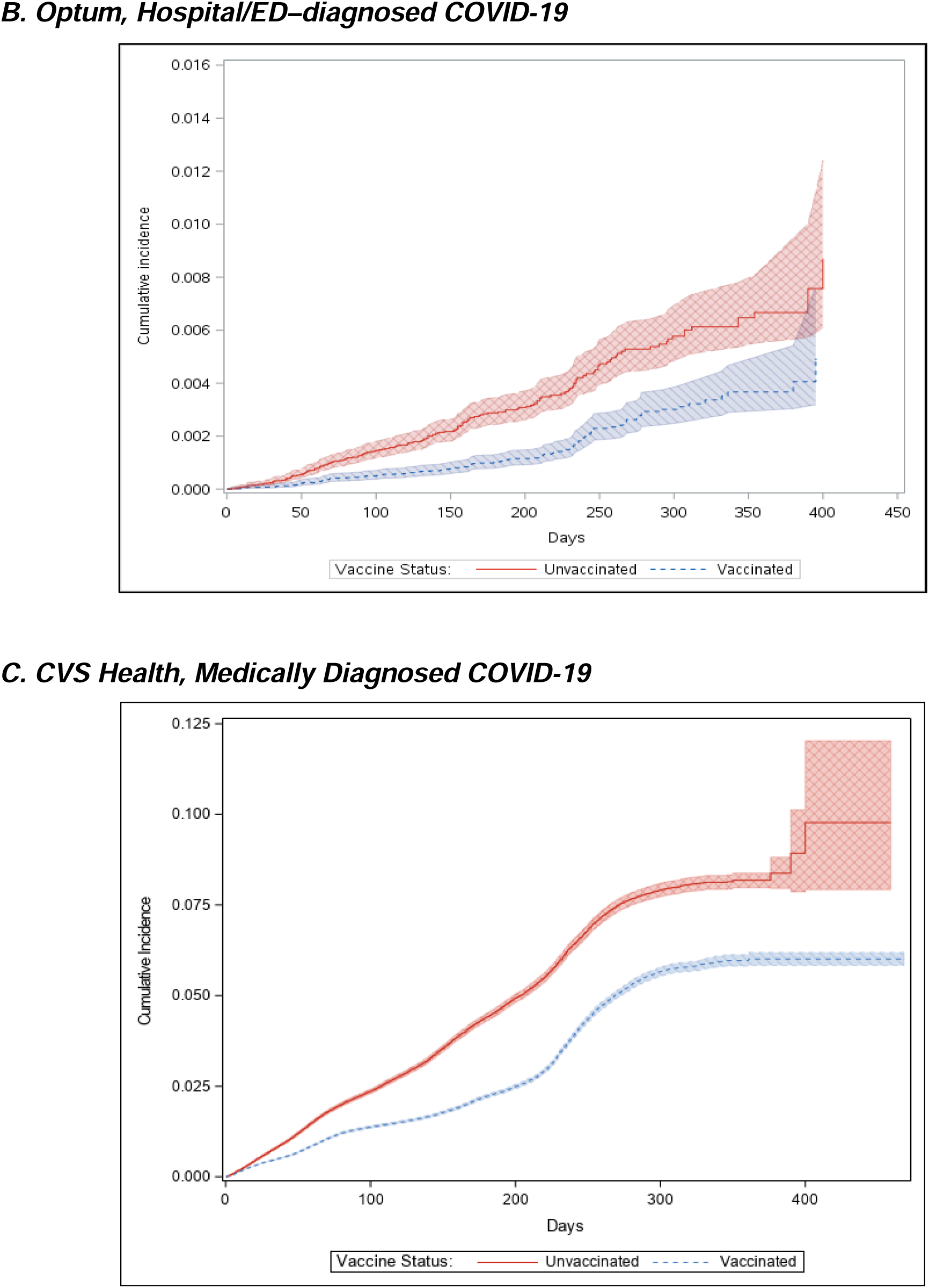
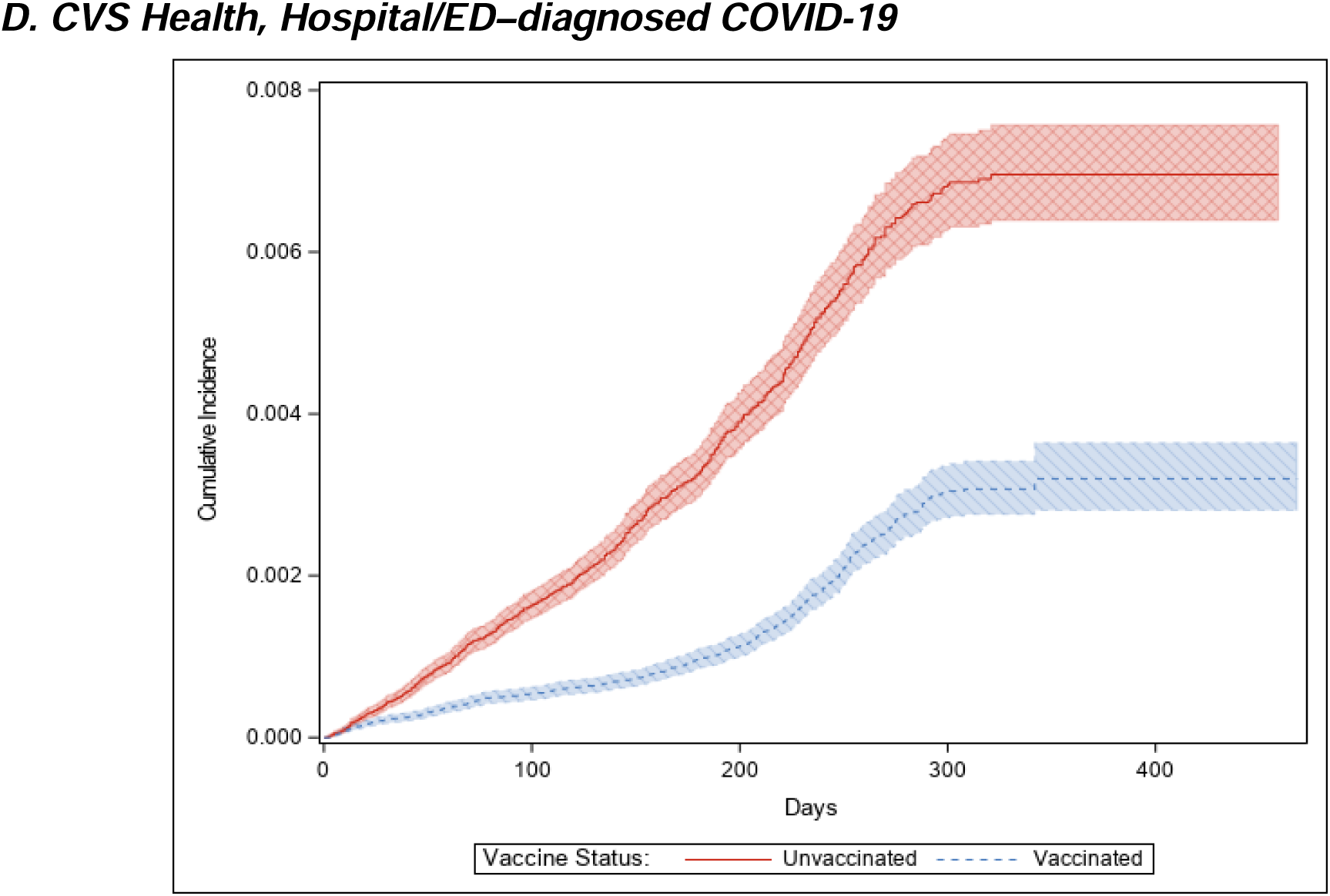
Weighted Cumulative Incidence of COVID-19 Outcomes in Children Aged 5–17 Years Receiving a Complete Primary Series of BNT162b2 COVID-19 Vaccine and Unvaccinated Children. COVID-19 = coronavirus disease 2019; ED = emergency department; sIPT = stabilized inverse probability of treatment weighting.

Children were eligible for inclusion on or after the date BNT162b2 was authorized for their age group (December 11, 2020, for ages 16–17; May 10, 2021, for ages 12–15; October 29, 2021, for ages 5–11). Vaccinated and matched unvaccinated children were required to meet the following inclusion criteria (eFigure 1): at least 365 days of continuous medical and pharmacy coverage before Time 0 (including the date of the age group–specific vaccine authorization to ensure observation of all COVID-19 vaccine doses); be aged within the vaccine-authorized age range at Time 0; and reside within the catchment area of the linked claims-IIS data. Children were excluded if they had a procedure or diagnostic code for one of the following before Time 0: monoclonal antibody or convalescent plasma treatment (90 days); COVID-19 diagnosis (30 days); fever, nausea/vomiting, rash diagnosis (3 days); hospitalization or ED visit (3 days); or hospitalization or long-term care residence (on Time 0). Children selected as unvaccinated comparators could subsequently be vaccinated and enter the vaccinated group with a new Time 0.

### Exposure Assessment

BNT162b2 doses were identified using brand-specific procedure codes for vaccine administration, pharmacy codes for vaccine products, or IIS vaccination records.^11,14,15^ Dose number was inferred from the chronological order of observed doses within a child’s record. An unbranded dose or a dose of the same brand occurring within 3 days following another dose was considered a duplicate and was removed; if a dose for a different brand was received within 3 days, the brand of the dose was considered unclassifiable. Follow-up began for all vaccinated children at dose 1 (Time 0) regardless of future vaccine dose receipt.

Children were followed from Time 0 until the study outcome or censoring at the first occurrence of the following: last day of the IIS-specific study period; disenrollment from health plan; or deviation from the vaccine exposure assigned at Time 0 (eFigure 2). For vaccinated children, deviation from vaccine exposure included receiving BNT162b2 dose 2 before day 17, failure to receive BNT162b2 dose 2 by day 42, receipt of any other brand of COVID-19 vaccine or an unclassifiable dose, or a third dose of BNT162b2 (eFigure 2). For unvaccinated children, deviation included receiving a first dose of any COVID-19 vaccine.

**Figure 2.**
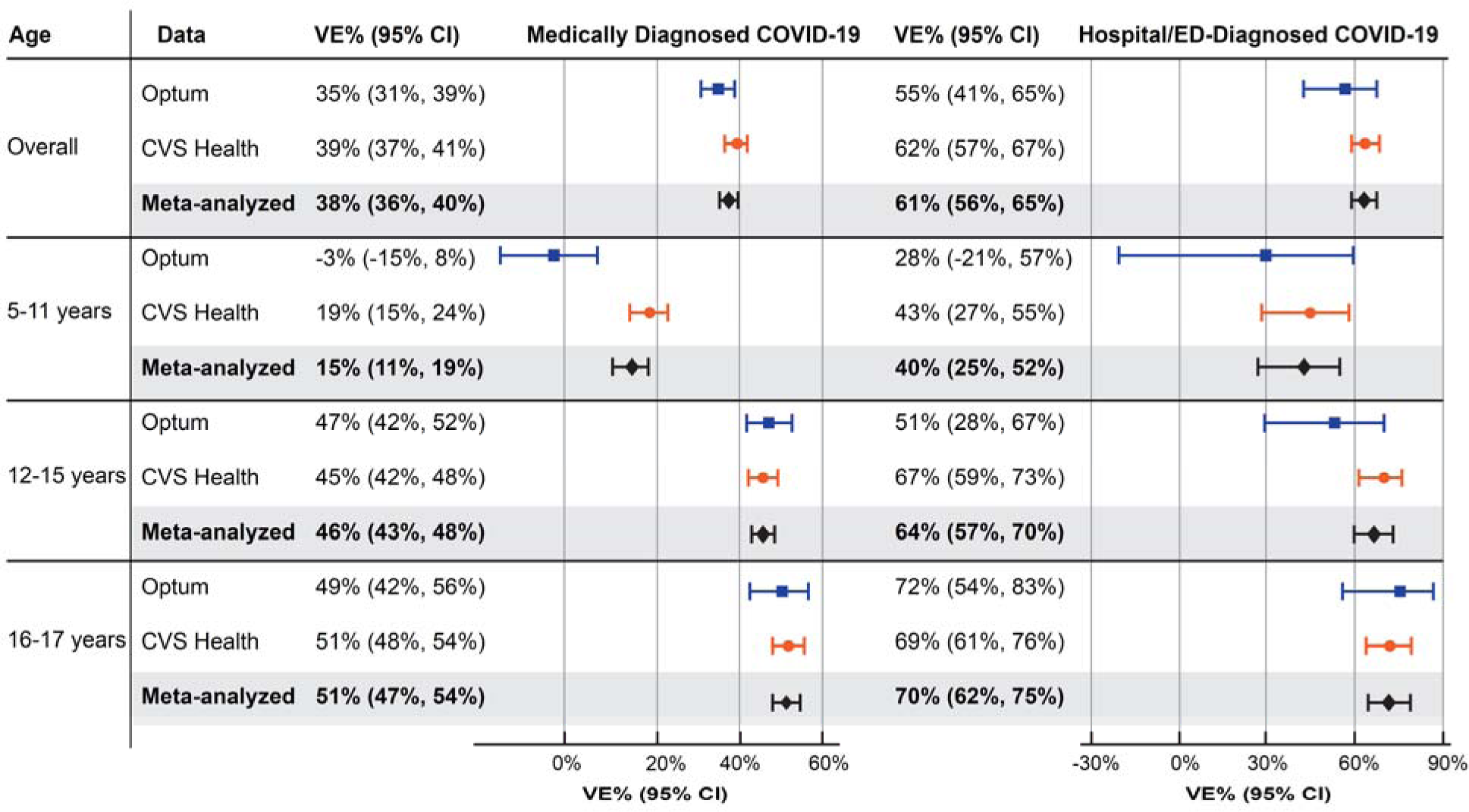
Estimated Effectiveness of Receiving a Complete Primary Series of BNT162b2 Compared With Being Unvaccinated in Children Aged 5–17 Years, Overall and by Age Group. CI = confidence interval; COVID-19 = coronavirus disease 2019; VE = vaccine effectiveness.

### Outcome Assessment

Two COVID-19 outcomes were evaluated separately: (1) medically diagnosed COVID-19, identified as a recorded COVID-19 diagnosis from hospital, ED, outpatient, or physician encounters; and (2) hospital/ED–diagnosed COVID-19. Recorded COVID-19 diagnosis codes (ICD-10-CM U07.1) were identified in claims in any coding position. The recorded date of the first diagnosis was assigned as the outcome date.

### Covariates

Descriptive characteristics were measured on or before Time 0 (eFigure 1) using enrollment, diagnosis, procedure, and pharmacy data.^16^ Covariates included demographics, comorbidities, frailty indicators, healthcare utilization, and conditions potentially increasing risk of severe COVID-19.^13^

### Statistical Analysis

Analyses were performed separately by data source. The distribution of characteristics by vaccination group were described with means, standard deviations (SD), medians, and first and third quartiles (Q1, Q3) for continuous variables, and counts and proportions for categorical variables. Covariate balance between vaccination groups was evaluated with absolute standardized differences.^17^

Propensity scores were estimated with multivariable logistic regression models including all prespecified covariates and matching factors. Stabilized inverse probability of treatment (sIPT) weights were estimated from propensity scores with truncation below the first percentile and above the 99th percentile of the propensity score distribution.

The cumulative incidence of each COVID-19 outcome was estimated in the sIPT-weighted vaccine exposure groups as 1 minus the Kaplan-Meier estimator.^18^ Hazard ratios (HRs) for the association of vaccination status with COVID-19 outcomes were estimated using sIPT-weighted Cox proportional hazards models; 95% confidence intervals (CIs) were estimated with robust sandwich variance estimators.^19^ Cumulative incidence and HRs in the first 14 days of follow-up were evaluated as a negative control outcome (COVID-19 vaccines are not expected to produce an immune response until 10-14 days after vaccination).^20,21^

Subgroup analyses were performed by age group (5–11, 12–15, 16–17 years) and by variant era (pre-delta era, December 11, 2020–May 31, 2021; delta era, June 1, 2021–December 24, 2021; omicron era, December 25, 2021–end of data availability^22^). Variant era analyses were restricted to children with Time 0s within the era, with follow-up censored on the last day of the era. A post-hoc analysis evaluated the distribution of person-time spent in each variant era by age subgroup resulting from the staggered authorizations by age group.

Quantitative bias analyses^23,24^ estimated the impact of potential misclassification because of missing vaccine records by estimating corrected HRs accounting for a range of vaccine exposure sensitivities (Supplemental Methods). A sensitivity analysis evaluated potential informative censoring by delaying censoring 7 days after receipt of a censoring vaccine dose. Additional sensitivity analyses evaluated the impact of potential outcome misclassification resulting from a recorded COVID-19 diagnosis on the same day as COVID-19 vaccination by removing Time 0 from follow-up and reordering censoring criteria so censoring for receipt of a censoring dose occurred first.

Data source–specific estimates were meta-analyzed using fixed-effects meta-analysis models. Analyses were performed with SAS version 9.4 (SAS Institute, Cary, NC). This surveillance activity was conducted as part of the FDA public health surveillance mandate and was not subject to Institutional Review Board oversight. The study protocol was publicly posted on the BEST Initiative website.^16^

## RESULTS

We identified 95,161 eligible children in Optum and 365,312 in CVS Health databases aged 5– 17 years who received a first dose of BNT162b2 during the study period. In Optum, 97% of vaccinated children could be exact matched to an unvaccinated child, leaving 92,338 in each vaccine exposure group (132,528 unique children); in CVS Health, 99% were matched, leaving 361,317 in each group (694,621 unique children) (eFigure 3). In both groups in both data sources, the mean age was 12 years (SD 4 years), and 50% were female. The largest proportion resided in the Midwest in Optum (47%), and in the West in CVS Health (42%). Characteristics of the groups were well balanced on all measured characteristics in both data sources (selected characteristics in Table 1; complete characteristics in eTable1). The propensity score distributions in the matched vaccinated and unvaccinated children were similar, suggesting comparability between the groups before weighting (eFigure 4).

**Figure 3.**
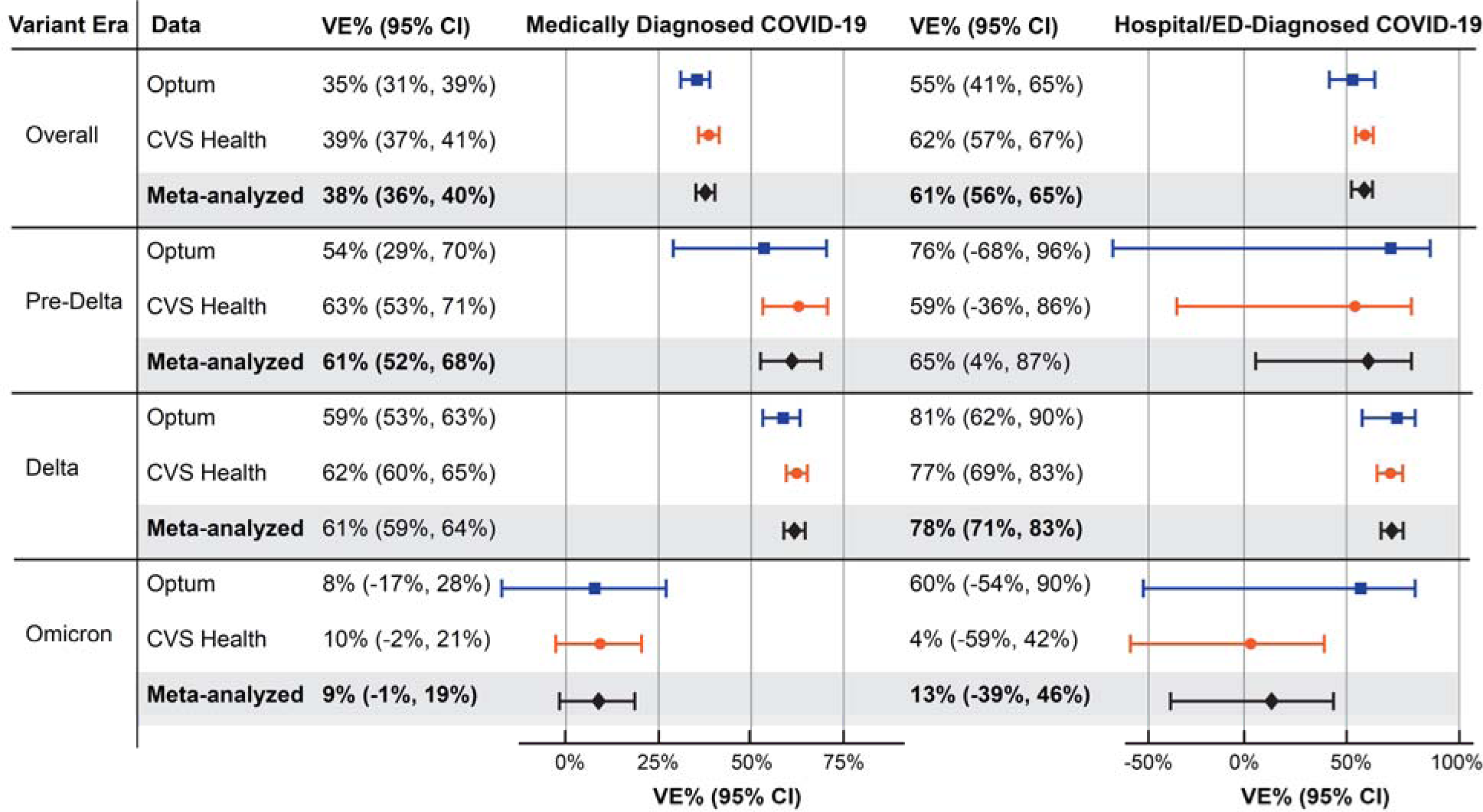
Estimated Effectiveness of Receiving a Complete Primary Series of BNT162b2 Compared With Being Unvaccinated in Children Aged 5–17 Years, Overall and by Variant Era. CI = confidence interval; COVID-19 = coronavirus disease 2019; VE = vaccine effectiveness. Note: pre-delta era, December 11, 2020–May 31, 2021; delta era, June 1, 2021–December 24, 2021; omicron era, December 25, 2021–end of data availability.

**Table 1.** Selected Characteristics of Children Vaccinated With BNT162b2 COVID-19 Vaccine and Matched Unvaccinated Children.

| Characteristic | Optum |  |  | CVS Health |  |  |
| --- | --- | --- | --- | --- | --- | --- |
|  | Vaccinated with BNT162b2<br>N = 92,338 | Matched<sup>a</sup><br>Unvaccinated<br>N = 92,338 | ASD | Vaccinated with BNT162b2<br>N = 361,317 | Matched<sup>a</sup><br>Unvaccinated<br>N = 361,317 | ASD |
| **Characteristics at Time 0** |  |  |  |  |  |  |
| Age, years |  |  |  |  |  |  |
| Median (Q1, Q3) | 13 (9, 15) | 13 (9, 15) |  | 12 (9, 15) | 12 (9, 15) | 0.00 |
| Mean (SD) | 12.11 (3.60) | 12.08 (3.64) | 0.01 | 11.76 (3.66) | 11.71 (3.72) | 0.01 |
| Sex, N (%) |  |  |  |  |  |  |
| Male | 46,516 (50.38%) | 46,516 (50.38%) | 0.00 | 181,416 (50.21%) | 181,416 (50.21%) | 0.00 |
| Female | 45,822 (49.62%) | 45,822 (49.62%) | 0.00 | 179,901 (49.79%) | 179,901 (49.79%) | 0.00 |
| Region, N (%) |  |  |  |  |  |  |
| Northeast | 13,051 (14.13%) | 13,051 (14.13%) | 0.00 | 64,684 (17.90%) | 64,684 (17.90%) | 0.00 |
| South | 12,959 (14.03%) | 12,959 (14.03%) | 0.00 | 69,202 (19.15%) | 69,202 (19.15%) | 0.00 |
| Midwest | 43,572 (47.19%) | 43,572 (47.19%) | 0.00 | 75,498 (20.90%) | 75,498 (20.90%) | 0.00 |
| West | 22,756 (24.64%) | 22,756 (24.64%) | 0.00 | 151,933 (42.05%) | 151,933 (42.05%) | 0.00 |
| **Characteristics in the 365 days before Time 0, N (%)** |  |  |  |  |  |  |
| Influenza vaccination | 47,759 (51.72%) | 47,759 (51.72%) | 0.00 | 187,076 (51.78%) | 187,076 (51.78%) | 0.00 |
| Pneumococcal vaccination | 87 (0.09%) | 77 (0.08%) | 0.00 | 360 (0.10%) | 368 (0.10%) | 0.00 |
| Well-check/well-child preventive health care visit | 63,290 (68.54%) | 62,515 (67.70%) | 0.02 | 253,176 (70.07%) | 246,607 (68.25%) | 0.04 |
| **Characteristics assessed using all available data at or before Time 0, N (%)** |  |  |  |  |  |  |
| Autoimmune disorders | 1,067 (1.16%) | 1,049 (1.14%) | 0.00 | 4,273 (1.18%) | 3,995 (1.11%) | 0.01 |
| Chronic lung diseases (e.g., asthma, COPD, cystic fibrosis, pulmonary embolism) | 10,086 (10.92%) | 10,081 (10.92%) | 0.00 | 43,858 (12.14%) | 44,283 (12.26%) | 0.00 |
| Dementia or other neurological conditions | 3,110 (3.37%) | 3,331 (3.61%) | 0.01 | 12,075 (3.34%) | 12,542 (3.47%) | 0.01 |
| Diabetes mellitus, type 1 or 2 | 436 (0.47%) | 416 (0.45%) | 0.00 | 1,480 (0.41%) | 1,336 (0.37%) | 0.01 |
| Heart conditions (e.g., heart failure, coronary artery disease, arrhythmias) | 3,037 (3.29%) | 3,135 (3.40%) | 0.01 | 12,537 (3.47%) | 13,118 (3.63%) | 0.01 |
| Immunocompromised state | 458 (0.50%) | 458 (0.50%) | 0.00 | 1,674 (0.46%) | 1,674 (0.46%) | 0.00 |
|  | Vaccinated with BNT162b2<br>N = 92,338 | Matched<sup>a</sup><br>Unvaccinated<br>N = 92,338 | ASD | Vaccinated with BNT162b2<br>N = 361,317 | Matched<sup>a</sup><br>Unvaccinated<br>N = 361,317 | ASD |
| Mental health conditions | 17,813 (19.29%) | 16,871 (18.27%) | 0.03 | 63,102 (17.46%) | 58,900 (16.30%) | 0.03 |
| Obese or severely obese | 7,750 (8.39%) | 8,138 (8.81%) | 0.01 | 35,224 (9.75%) | 37,775 (10.45%) | 0.02 |
| At least one COVID-19 laboratory performed | 36,002 (38.99%) | 32,568 (35.27%) | 0.08 | 174,485 (48.29%) | 157,714 (43.65%) | 0.09 |
| COVID-19 diagnoses occurring outside of a hospital or emergency department | 2,759 (2.99%) | 2,751 (2.98%) | 0.00 | 10,878 (3.01%) | 10,812 (2.99%) | 0.00 |
| Hospitalization or emergency department-diagnosed COVID-19 | 116 (0.13%) | 129 (0.14%) | 0.00 | 514 (0.14%) | 643 (0.18%) | 0.01 |

ASD = absolute standardized difference; COPD = Chronic obstructive pulmonary disease; COVID-19 = coronavirus disease 2019; Q1, Q3 = first and third quartiles; SD = standard deviation; US = United States.
<sup>a</sup> Matched children on calendar date, age group, sex, US county of residence, immunocompromised status, pregnancy status, history of COVID-19 diagnosis, presence of a comorbidity identified by the Centers for Disease Control and Prevention as increasing individuals' risk of severe COVID-19, influenza vaccination in previous year.

The amount of follow-up time ranged across data sources and analyses, but the maximum follow-up time was 529 days in Optum and 468 days in CVS Health; the median follow-up tended to be longer for the vaccinated group in both data sources for each analysis (eTable 2). The cumulative incidence of COVID-19 outcomes over time by vaccination group is shown in Figure 1. Across all analyses in both data sources, the rates of hospital/ED-diagnosed COVID-19 cases were relatively small (Optum, 41.2 per 10,000 person-years; CVS Health, 44.1 per 10,000 person-years). For the main analyses, the risk of COVID-19 outcomes was higher in the unvaccinated than vaccinated groups throughout follow-up. The overall sIPT-weighted VE estimates for medically diagnosed COVID-19 were 35% (95% CI, 31%-39%) in Optum and 39% (95% CI, 37%-41%) in CVS Health (meta-analyzed VE = 38% [95% CI, 36%-40%]). For hospital/ED-diagnosed COVID-19, VE estimates were 55% (95% CI, 41%-65%) in Optum and 62% (95% CI, 57%-67%) in CVS Health (meta-analyzed VE = 61% [95% CI, 56%-65%]) (Figure 2, eTable 3). Using sensitivity estimates for the claims/IIS-based measures of COVID-19 vaccine exposure of 83% and 71% in Optum and 89% and 69% in CVS Health, quantitative bias analyses suggested that the observed VE estimates may underestimate the true VE by 2% to 13% (eTable 4).

During the 14-day negative control period (Time 0 to day 13), the absolute risk of COVID-19 outcomes was low, and the absolute difference between the vaccinated and unvaccinated groups was small (eFigure 5). However, estimated VEs during this period indicated potential associations of vaccination with medically diagnosed COVID-19: Optum VE = 15% (95% CI, - 1.7% to 28%); CVS Health VE = 25% (95% CI, 17%-32%) (eTable 5); negative control estimates for hospital/ED–diagnosed COVID-19 were imprecise as a result of having few cases. Post-hoc explorations of COVID-19 testing patterns during the negative control period in Optum suggested outcome misclassification caused by less frequent COVID-19 testing and diagnoses in the vaccinated group in the 3 to 4 days after vaccination (eFigure 7).

When stratifying by age subgroup, VE estimates were generally high and similar across data sources for 16- to 17-year-olds (medically diagnosed COVID-19 meta-analyzed VE = 51% [95% CI, 47%-54%]; hospital/ED–diagnosed COVID-19 meta-analyzed VE = 70% [95% CI, 62%-75%]) and 12- to 15-year-olds (medically diagnosed COVID-19 meta-analyzed VE = 46% [95% CI, 43%-48%]; hospital/ED–diagnosed COVID-19 meta-analyzed VE = 64% [95% CI, 57%-70%]) (Figure 2). In both data sources, VE estimates were lower in 5- to 11-year-olds. For 5- to 11-year-olds, the meta-analyzed VE for hospital/ED–diagnosed COVID-19 was 40% (95% CI, 25%-52%); however, for medically diagnosed COVID-19, heterogeneity was observed across data sources (*p* < 0.01), with an Optum VE estimate of −3% (95% CI, −15% to 8%) and a CVS Health VE estimate of 19% (95% CI, 15%-24%).

When evaluating VE by variant era, the largest sample sizes and follow-up were observed in the delta era (eTable 6, eTable 7). Variant era was highly correlated with age group, with the majority of person-time in the pre-delta era coming from the 16- to 17-year-olds, the majority of delta era person-time from 12- to 15-year-olds, and the majority of omicron era person-time coming from 5- to 11-year-olds (eTable 6). The VE estimates during the omicron era against medically diagnosed COVID-19 (meta-analyzed VE = 9% [95% CI, −1% to 19%]) and against hospital/ED–diagnosed COVID-19 (meta-analyzed VE = 13% [95% CI, −39% to 46%]) were markedly lower than during the pre-delta or delta eras (Figure 3). However, although there was no statistical heterogeneity between the source-specific omicron era VE estimates against hospital/ED-diagnosed COVID-19 (*p* = 0.24), there was a large range between the VE estimates: Optum VE = 60% [95% CI, −54% to 90%]); CVS Health VE = 4% [95% CI, −59% to 42%]) (eTable 7). Results of sensitivity analyses were similar to those of the primary analyses (eFigure 6).

## DISCUSSION

In this large, real-world evaluation of the effectiveness of monovalent BNT162b2 vaccination in children aged 5–17 years, lower rates of COVID-19 diagnoses were observed among children receiving a complete primary series of BNT162b2 compared with unvaccinated children, indicating that this vaccine is effective in routine care. Vaccine effectiveness was higher for hospital/ED–diagnosed COVID-19 than for any medically diagnosed COVID-19 and lower for children aged 5–11 years and during the omicron era.

These observed VE estimates in children are generally lower than many VE estimates reported for the primary series of BNT162b2 in adults.^25–27^ US children aged less than 16 were vaccinated relatively late in the pandemic after many months of potential COVID-19 exposures and infection, largely during the delta variant era. Many COVID-19 infections in children were relatively mild during the early pandemic,^28,29^ and previous history of COVID-19 infection earlier in the pandemic before vaccination may have conveyed some level of natural immunity in both the vaccinated and unvaccinated groups, reducing the observed VE estimates.

Our study utilized two data sources, and the overall results of higher VE estimates for hospital/ED–diagnosed COVID-19 than for medically diagnosed COVID-19 across all age groups is consistent with the results of other studies.^30,31^ Additionally, many of the age and variant subgroup results were largely consistent across data sources. Children aged 5–11 years were the last age group receiving vaccine authorization during the study period, making it difficult to disentangle the effect of age group and variant era. Both data sources suggested lower VE in 5- to 11-year-olds and in the omicron era—consistent with other studies^31^—but there were key differences in the magnitude of the VE estimates across data sources in these subgroups. Although calendar time and geography were balanced across vaccination groups within each data source (accounting for local differences in COVID-19 circulation and severity), the two data sources covered different geographic areas and timeframes. For example, the end date for all IIS jurisdictions in CVS Health was March 31, 2022, but in Optum, the end dates varied from September 30, 2021 (before the beginning of the omicron era) to May 31, 2022 (into the “second omicron wave” starting in April/May 2022).^32^

This study has several strengths including a large sample size, inclusion of multiple US geographic regions, and combining vaccine administrative claims with IIS vaccine records. The linkage of claims to IIS data supplemented the vaccine exposure data and reduced vaccine exposure misclassification from vaccine doses not recorded in claims data. However, some vaccine administrations may still have been missed. The study used external estimates of vaccine coverage among individuals younger than 65 years to quantify potential residual exposure misclassification and applied quantitative bias analysis to correct VE estimates. Because younger children tended to be vaccinated later in the study period when mass vaccination clinics were less common, and younger children may have lower levels of vaccination compared with adults,^33^ the extent of exposure misclassification may be overestimated.

Because pandemic conditions varied widely across geographic areas and time periods, vaccinated and unvaccinated children were matched on calendar time and county of residence to account for these differences. The eligibility and matching criteria were designed to identify vaccinated and unvaccinated children who were eligible for vaccination on each calendar day, avoiding selection bias. Starting follow-up on Time 0 without considering future vaccination behaviors avoided immortal person-time bias.^34^

This real-world study has limitations. This study did not analyze laboratory-confirmed COVID-19 status, so the study relied on recorded claims-based diagnoses of COVID-19. Although COVID-19 diagnosis codes have shown reasonable validity for hospitalized cases,^35–40^ many COVID-19 cases may never be formally diagnosed in a health care setting, and the dynamics of COVID-19 testing and diagnosis changed over time.

Despite matching and propensity score weighting, residual and unmeasured confounding may remain. The negative control analysis suggested a potential difference between the exposure groups immediately after vaccination when vaccines are assumed to have no biologic effect. However, the post hoc negative control analysis demonstrated differential COVID-19 testing and diagnoses in the 3-4 days after Time 0, because recently vaccinated individuals may not seek COVID-19 testing.^41^ This difference in testing and diagnosis behavior appeared to resolve after day 4, but longer term differences in health care–seeking behavior cannot be ruled out.

Because of the staged authorization of vaccines by age group, the study could not evaluate vaccine effectiveness by variant era and age groups simultaneously (e.g., only children aged 16-17 years were authorized to be vaccinated until nearly the end of the pre-delta era).

## CONCLUSIONS

Receiving a complete primary series of BNT162b2 was associated with reduced COVID-19 incidence compared to being unvaccinated in the pediatric population. BNT162b2’s effectiveness was higher for hospital/ED-diagnosed COVID-19 than for any medically diagnosed COVID-19, and it was higher among children aged 12-17 compared to 5-11 years. In the rapidly changing dynamics of the COVID-19 pandemic, additional real-world studies are needed to evaluate COVID-19 vaccine effectiveness as booster doses and additional vaccine brands become available for this population.

## Supporting information

eTable

## Data Availability

The data that has been used is confidential.

## ACKNOWLEDGEMENTS

The authors acknowledge the assistance of the following: Shanlai Shangguan, MPH, Blair Cha, BA, Wenxuan Zhou, BS, and Jessica Hervol, MPH, of Acumen, LLC for editorial review of the manuscript and interpretation of results; Anne Marie Kline, MS, Nancy B. Shaik, BS, Ana M. Martinez-Baquero, MA, Vaibhav Sharma, MS, and Smita Bhatia, MCA, of CVS Health Clinical Trial Services for acquisition and analysis of the data; Vivian Wilt, MPH, Julia McIlmail, BS, and Rebecca Warsawski, MPH, of Optum Epidemiology for project management; Michael Kirksey, MBA, of Optum Enterprise Analytics for data analysis; Karen Schneider, PhD, and Emily Myers, MPH, of Optum Serve Consulting for feasibility evaluation and analytic support; Eli Wolter and Edward Novak, MS, of Optum Technology for data acquisition, processing, and technology leadership; Rebecca Braun, BA, of Optum Serve Consulting for data and information technology support; Megan Ketchell, BS, of Optum Serve Consulting for project coordination and communications support; Kathryn Federici, MSW, of Optum Serve Consulting for communications and legal support; Sarah Harris, MA, of RTI International for project management support; Michelle Myers, BS, and Virginia Ferguson, BFA, of RTI International for editorial review of the manuscript; and Melissa McPheeters, PhD, of RTI International for supervision.

