## Supplementary material for "Effectiveness of BNT162b2 COVID-19 Vaccination in Children Aged 5–17 Years in the United States": eTable

* Co-first authors

Corresponding Author:

J. Bradley Layton

### SUPPLEMENTAL INFORMATION

#### Supplemental Methods: Data Sources

The Optum database includes enrollment, adjudicated prescription drug claims, and preadjudicated hospital and physician health insurance claims. The preadjudicated claims database includes claims for privately insured and Medicare Advantage enrollees. Hospital and physician claims undergo initial processing daily from a large number of providers across the United States who accept patients with health insurance. Optum has established an ongoing weekly update schedule to incorporate newly processed claims into the preadjudicated claims database. This data source was utilized to reduce the delay between the occurrence of healthcare services and their presence in the database. The preadjudicated claims have an approximately 2-month delay for 90% completeness for inpatient claims and over 70% completeness at 1 month for outpatient claims. To enhance vaccine administration capture, the Optum claims were linked to COVID-19 vaccination records from 10 Immunization Information Systems (IIS).^1,2^

The CVS Health data include Aetna enrollment, dispensed prescription drug, and adjudicated hospital and physician health insurance claims. The Aetna adjudicated claims database includes claims for Aetna commercially insured and Aetna Medicare Advantage enrollees. The adjudicated claims are 80% complete for inpatient claims at 3 months and outpatient claims at 2 months. To enhance vaccine administration capture, the CVS Health/Aetna claims were linked to COVID-19 vaccination records from 11 IIS.

#### Supplemental Methods: Quantitative Bias Analysis

A simple quantitative bias analysis evaluating the impact of exposure misclassification was performed. Estimates of statewide receipt of at least one COVID-19 vaccine dose from the Centers for Disease Control and Prevention (CDC), state departments of health, and capture-recapture methods^3-5^ were compared with observed state-level estimates in the linked claims-IIS data sources to estimate potential sensitivities of our measure of vaccine exposure.

The primary analyses estimated hazard ratios, but for the purpose of the quantitative bias analyses, risk ratios (RR) and 95% confidence intervals (CIs) were estimated in the weighted cohorts using a fixed 61-day follow-up time for both outcomes.

Specificity of the study’s vaccine assessment is assumed to be 100% (e.g., all observed claims or IIS records are assumed to be true vaccination events, and no truly unvaccinated children was misclassified as being vaccinated). Estimates of underrecording were calculated by comparing each jurisdiction’s vaccine coverage estimates for the population aged 65 years or less to age standardized estimates from CDC, state departments of health, and a mathematically derived vaccine estimate using capture-recapture methods^4^ with IIS and claims as the two sources. Using these estimates of underrecording, two bounds of sensitivity were estimated at 71% and 83% in Optum and 89% and 69% in CVS Health. “Corrected” RR estimates were generated for each outcome by reassigning exposure status from unvaccinated to vaccinated based on the two sensitivity estimates. A correction factor was then estimated as follows:

$$bias correction factor=1-\frac{corrected RR}{uncorrected RR}$$

The bias correction factors were then applied to the observed VE estimates from the primary analyses, generating an estimate of VE corrected for exposure misclassification.

eTable 1.	Complete Characteristics of Children Vaccinated With BNT162b2 COVID‑19 Vaccine and Matched Unvaccinated Children

##### A. Optum

| Characteristic | Vaccinated N = 92,338 | Unvaccinated N = 92,338 | ASD |
| --- | --- | --- | --- |
| Characteristics assessed at Time 0 |  |  |  |
| Age, years |  |  |  |
| Median (Q1, Q3) | 13 (9, 15) | 13 (9, 15) |  |
| Mean (SD) | 12.11 (3.60) | 12.08 (3.64) | 0.01 |
| Sex, N (%) |  |  |  |
| Male | 46,516 (50.38%) | 46,516 (50.38%) | 0.00 |
| Female | 45,822 (49.62%) | 45,822 (49.62%) | 0.00 |
| Region, N (%) |  |  |  |
| Northeast | 13,051 (14.13%) | 13,051 (14.13%) | 0.00 |
| South | 12,959 (14.03%) | 12,959 (14.03%) | 0.00 |
| Midwest | 43,572 (47.19%) | 43,572 (47.19%) | 0.00 |
| West | 22,756 (24.64%) | 22,756 (24.64%) | 0.00 |
| Pregnant at Time 0, N (%) | < 11 | < 11 | 0.00 |
| Characteristics in the 365 days before Time 0, N (%) |  |  |  |
| Hospitalizations |  |  |  |
| 0 | 78,015 (84.49%) | 77,519 (83.95%) | 0.01 |
| 1 | 9,757 (10.57%) | 10,098 (10.94%) | 0.01 |
| 2+ | 4,566 (4.94%) | 4,721 (5.11%) | 0.01 |
| Emergency department visits |  |  |  |
| 0 | 85,807 (92.93%) | 85,029 (92.08%) | 0.03 |
| 1 | 5,665 (6.14%) | 6,254 (6.77%) | 0.03 |
| 2+ | 866 (0.94%) | 1,055 (1.14%) | 0.02 |
| Skilled nursing facility stay | 11 (0.01%) | < 11 | 0.00 |
| Influenza vaccination | 47,759 (51.72%) | 47,759 (51.72%) | 0.00 |
| Pneumococcal vaccination | 87 (0.09%) | 77 (0.08%) | 0.00 |
| Encounter for cancer screening | 194 (0.21%) | 220 (0.24%) | 0.01 |
| Eye examination | 20,534 (22.24%) | 20,462 (22.16%) | 0.00 |
| Colonoscopy | 114 (0.12%) | 114 (0.12%) | 0.00 |
| Bone mineral density test | 57 (0.06%) | 54 (0.06%) | 0.00 |
| Well-check/well-child preventive healthcare visit | 63,290 (68.54%) | 62,515 (67.70%) | 0.02 |
| Arthritis | 6,683 (7.24%) | 6,943 (7.52%) | 0.01 |
| Lipid abnormality | 787 (0.85%) | 795 (0.86%) | 0.00 |
| Ambulance use or life support services | 856 (0.93%) | 956 (1.04%) | 0.01 |
| Weakness | 1,126 (1.22%) | 1,127 (1.22%) | 0.00 |
| Pregnancy completion before Time 0 | 15 (0.03%) | 39 (0.09%) | 0.02 |
| Characteristics assessed using all available data, N (%) |  |  |  |
| Autoimmune disorders | 1,067 (1.16%) | 1,049 (1.14%) | 0.00 |
| Cancer | 193 (0.21%) | 213 (0.23%) | 0.00 |
| Chronic kidney disease or renal disease | 194 (0.21%) | 185 (0.20%) | 0.00 |
| Chronic liver disease | 209 (0.23%) | 205 (0.22%) | 0.00 |
| Chronic lung diseases (e.g., asthma, COPD, cystic fibrosis, pulmonary embolism) | 10,086 (10.92%) | 10,081 (10.92%) | 0.00 |
| Dementia or other neurological conditions | 3,110 (3.37%) | 3,331 (3.61%) | 0.01 |
| Diabetes mellitus, type 1 or 2 | 436 (0.47%) | 416 (0.45%) | 0.00 |
| Down syndrome | 113 (0.12%) | 119 (0.13%) | 0.00 |
| Heart conditions (e.g., heart failure, coronary artery disease, arrhythmias) | 3,037 (3.29%) | 3,135 (3.40%) | 0.01 |
| Hypertension | 389 (0.42%) | 413 (0.45%) | 0.00 |
| Immunocompromised state | 458 (0.50%) | 458 (0.50%) | 0.00 |
| Mental health conditions | 17,813 (19.29%) | 16,871 (18.27%) | 0.03 |
| Obese or severely obese | 7,750 (8.39%) | 8,138 (8.81%) | 0.01 |
| Sickle cell disease or thalassemia | 142 (0.15%) | 137 (0.15%) | 0.00 |
| Stroke or cerebrovascular disease | 97 (0.11%) | 97 (0.11%) | 0.00 |
| Tuberculosis | < 11 | < 11 | 0.00 |
| At least one COVID-19 laboratory performed | 36,002 (38.99%) | 32,568 (35.27%) | 0.08 |
| COVID-19 diagnoses occurring outside of a hospital or emergency department | 2,759 (2.99%) | 2,751 (2.98%) | 0.00 |
| Hospitalization or emergency department-diagnosed COVID-19 | 116 (0.13%) | 129 (0.14%) | 0.00 |

ASD = absolute standardized difference; NA = not applicable; Q1 = first quartile; Q3 = third quartile; SD = standard deviation.

Note: Optum privacy rules require masking cell sizes < 11 individuals.

##### B. CVS Health

| Characteristic | Vaccinated N = 361,317 | Unvaccinated N = 361,317 | ASD |
| --- | --- | --- | --- |
| Characteristics assessed at Time 0 |  |  |  |
| Age, years |  |  |  |
| Median (Q1, Q3) | 12 (9, 15) | 12 (9, 15) | 0.00 |
| Mean (SD) | 11.76 (3.66) | 11.71 (3.72) | 0.01 |
| Sex, N (%) |  |  |  |
| Male | 181,416 (50.21%) | 181,416 (50.21%) | 0.00 |
| Female | 179,901 (49.79%) | 179,901 (49.79%) | 0.00 |
| Region, N (%) |  |  |  |
| Northeast | 64,684 (17.90%) | 64,684 (17.90%) | 0.00 |
| South | 69,202 (19.15%) | 69,202 (19.15%) | 0.00 |
| Midwest | 75,498 (20.90%) | 75,498 (20.90%) | 0.00 |
| West | 151,933 (42.05%) | 151,933 (42.05%) | 0.00 |
| Pregnant at Time 0, N (%) | 4 (0.00%) | 4 (0.00%) | 0.00 |
| Characteristics in the 365 days before Time 0, N (%) |  |  |  |
| Hospitalizations |  |  |  |
| 0 | 310,325 (85.89%) | 308,799 (85.46%) | 0.01 |
| 1 | 34,669 (9.60%) | 36,038 (9.97%) | 0.01 |
| 2+ | 16,323 (4.52%) | 16,480 (4.56%) | 0.00 |
| Emergency department visits |  |  |  |
| 0 | 337,110 (93.30%) | 333,776 (92.38%) | 0.04 |
| 1 | 21,207 (5.87%) | 23,725 (6.57%) | 0.03 |
| 2+ | 3,000 (0.83%) | 3,816 (1.06%) | 0.02 |
| Skilled nursing facility stay | 47 (0.01%) | 36 (0.01%) | 0.00 |
| Influenza vaccination | 187,076 (51.78%) | 187,076 (51.78%) | 0.00 |
| Pneumococcal vaccination | 360 (0.10%) | 368 (0.10%) | 0.00 |
| Encounter for cancer screening | 676 (0.19%) | 610 (0.17%) | 0.00 |
| Eye examination | 117,681 (32.57%) | 114,902 (31.80%) | 0.02 |
| Colonoscopy | 463 (0.13%) | 430 (0.12%) | 0.00 |
| Bone mineral density test | 256 (0.07%) | 196 (0.05%) | 0.01 |
| Well-check/well-child preventive healthcare visit | 253,176 (70.07%) | 246,607 (68.25%) | 0.04 |
| Arthritis | 23,131 (6.40%) | 23,328 (6.46%) | 0.00 |
| Lipid abnormality | 4,352 (1.20%) | 3,936 (1.09%) | 0.01 |
| Ambulance use or life support services | 3,014 (0.83%) | 3,554 (0.98%) | 0.02 |
| Weakness | 3,678 (1.02%) | 3,480 (0.96%) | 0.01 |
| Pregnancy completion before Time 0 for females | 56 (0.02%) | 114 (0.03%) | 0.01 |
| Characteristics assessed using all available data, N (%) |  |  |  |
| Autoimmune disorders | 4,273 (1.18%) | 3,995 (1.11%) | 0.01 |
| Cancer | 800 (0.22%) | 907 (0.25%) | 0.01 |
| Chronic kidney disease or renal disease | 1,065 (0.29%) | 1,149 (0.32%) | 0.00 |
| Chronic liver disease | 730 (0.20%) | 747 (0.21%) | 0.00 |
| Chronic lung diseases (e.g., asthma, COPD, cystic fibrosis, pulmonary embolism) | 43,858 (12.14%) | 44,283 (12.26%) | 0.00 |
| Dementia or other neurological conditions | 12,075 (3.34%) | 12,542 (3.47%) | 0.01 |
| Diabetes mellitus, type 1 or 2 | 1,480 (0.41%) | 1,336 (0.37%) | 0.01 |
| Down syndrome | 396 (0.11%) | 386 (0.11%) | 0.00 |
| Heart conditions (e.g., heart failure, coronary artery disease, arrhythmias) | 12,537 (3.47%) | 13,118 (3.63%) | 0.01 |
| Hypertension | 1,531 (0.42%) | 1,568 (0.43%) | 0.00 |
| Immunocompromised state | 1,674 (0.46%) | 1,674 (0.46%) | 0.00 |
| Mental health conditions | 63,102 (17.46%) | 58,900 (16.30%) | 0.03 |
| Obese or severely obese | 35,224 (9.75%) | 37,775 (10.45%) | 0.02 |
| Sickle cell disease or thalassemia | 712 (0.20%) | 773 (0.21%) | 0.00 |
| Stroke or cerebrovascular disease | 509 (0.14%) | 597 (0.17%) | 0.01 |
| Tuberculosis | 58 (0.02%) | 44 (0.01%) | 0.00 |
| At least one COVID-19 laboratory performed | 174,485 (48.29%) | 157,714 (43.65%) | 0.09 |
| COVID-19 diagnoses occurring outside of a hospital or emergency department | 10,878 (3.01%) | 10,812 (2.99%) | 0.00 |
| Hospitalization or emergency department-diagnosed COVID-19 | 514 (0.14%) | 643 (0.18%) | 0.01 |

ASD = absolute standardized difference; Q1 = first quartile; Q3 = third quartile; SD = standard deviation.

eTable 2.	Distribution of Follow-up Person-time by Data Source, Vaccination Group, and Analysis

| COVID-19 Outcome | Vaccine Exposure Group | N | Person-time (Days) | | | |
| --- | --- | --- | --- | --- | --- | --- |
|  |  |  | Sum | Mean (SD) | Median (Q1, Q3) | Min, Max |
| **Optum** |  |  |  |  |  |  |
| Medically diagnosed | BNT162b2 | 92,338 | 15,334,736 | 166 (103) | 169 (58, 240) | 1, 512 |
|  | Unvaccinated | 92,338 | 11,114,593 | 120 (107) | 93 (29, 191) | 1, 503 |
| Hospitalization/ED-diagnosed | BNT162b2 | 92,338 | 15,566,203 | 169 (104) | 174 (62, 241) | 1, 512 |
|  | Unvaccinated | 92,338 | 11,400,524 | 123 (109) | 97 (30, 194) | 1, 529 |
| **CVS Health** |  |  |  |  |  |  |
| Medically diagnosed | BNT162b2 | 361,317 | 63,030,324 | 174 (92.48) | 146 (107, 249) | 1, 468 |
|  | Unvaccinated | 361,317 | 40,140,782 | 111 (100.3) | 84 (25, 147) | 1, 459 |
| Hospitalization/ED-diagnosed | BNT162b2 | 361,317 | 63,794,102 | 177 (92.51) | 146 (111, 251) | 1, 468 |
|  | Unvaccinated | 361,317 | 41,115,760 | 114 (101.82) | 87 (26, 153) | 1, 459 |

COVID-19 = coronavirus disease 2019; ED = emergency department; Min, Max = minimum, maximum; Q1, Q3 = first and third quartiles; SD = standard deviation.

eTable 3.	Estimated Effectiveness of Receiving a Complete Primary Series of BNT162b2 COVID‑19 Vaccine Compared to Being Unvaccinated in Children Aged 5–17 Years, Overall and in Age-specific Subgroups

##### A. Optum

| COVID-19 Outcome | Vaccine Exposure Group | N | Number of Events | Person-Time (Days) | sIPTW HR (95% CI) | sIPTW VE (95% CI) |
| --- | --- | --- | --- | --- | --- | --- |
| **Overall, ages 5–17 years** |  |  |  |  |  |  |
| Medically diagnosed | BNT162b2 | 92,338 | 2,629 | 15,334,736 | 0.65 (0.61, 0.69) | 35% (31%, 39%) |
|  | Unvaccinated | 92,338 | 2,792 | 11,114,593 | — | — |
| Hospitalization/ED-diagnosed | BNT162b2 | 92,338 | 118 | 15,566,203 | 0.45 (0.35, 0.59) | 55% (41%, 65%) |
|  | Unvaccinated | 92,338 | 186 | 11,400,524 | — | — |
| **Ages 5–11 years** |  |  |  |  |  |  |
| Medically diagnosed | BNT162b2 | 33,423 | 1,002 | 3,746,047 | 1.03 (0.92, 1.15) | -2.8% (-15%, 7.7%) |
|  | Unvaccinated | 33,423 | 764 | 2,950,100 | — | — |
| Hospitalization/ED-diagnosed | BNT162b2 | 33,423 | 34 | 3,828,835 | 0.72 (0.43, 1.21) | 28% (-21%, 57%) |
|  | Unvaccinated | 33,423 | 37 | 3,014,593 | — | — |
| **Ages 12–15 years** |  |  |  |  |  |  |
| Medically diagnosed | BNT162b2 | 39,560 | 1,090 | 7,556,338 | 0.53 (0.48, 0.58) | 47% (42%, 52%) |
|  | Unvaccinated | 39,560 | 1,322 | 5,274,074 | — | — |
| Hospitalization/ED-diagnosed | BNT162b2 | 39,560 | 54 | 7,654,311 | 0.49 (0.33, 0.72) | 51% (28%, 67%) |
|  | Unvaccinated | 39,560 | 76 | 5,422,148 | — | — |
| **Ages 16–17 years** |  |  |  |  |  |  |
| Medically diagnosed | BNT162b2 | 19,355 | 537 | 4,032,351 | 0.51 (0.44, 0.58) | 49% (42%, 56%) |
|  | Unvaccinated | 19,355 | 706 | 2,890,419 | — | — |
| Hospitalization/ED-diagnosed | BNT162b2 | 19,355 | 30 | 4,083,057 | 0.28 (0.17, 0.46) | 72% (54%, 83%) |
|  | Unvaccinated | 19,355 | 73 | 2,963,783 | — | — |

— indicates the reference group; CI = confidence interval; COVID‑19 = coronavirus disease 2019; ED = emergency department; HR = hazard ratio; sIPTW = stabilized inverse probability of treatment weighted; VE = vaccine effectiveness.

##### B. CVS Health

| COVID-19 Outcome | Vaccine Exposure Group | N | Number of Events | Person-Time (Days) | sIPTW HR (95% CI) | sIPTW VE (95% CI) |
| --- | --- | --- | --- | --- | --- | --- |
| **Overall, ages 5–17 years** |  |  |  |  |  |  |
| Medically diagnosed | BNT162b2 | 361,317 | 10,139 | 63,030,324 | 0.61 (0.59, 0.63) | 39% (37%, 41%) |
|  | Unvaccinated | 361,317 | 10,080 | 40,140,782 | — | — |
| Hospitalization/ED-diagnosed | BNT162b2 | 361,317 | 477 | 63,794,102 | 0.38 (0.33, 0.43) | 62% (57%, 67%) |
|  | Unvaccinated | 361,317 | 791 | 41,115,760 | — | — |
| **Ages 5–11 years** |  |  |  |  |  |  |
| Medically diagnosed | BNT162b2 | 152,460 | 3,650 | 16,280,047 | 0.81 (0.76, 0.85) | 19% (15%, 24%) |
|  | Unvaccinated | 152,460 | 3,135 | 10,846,098 | — | — |
| Hospitalization/ED-diagnosed | BNT162b2 | 152,460 | 144 | 16,513,481 | 0.57 (0.45, 0.73) | 43% (27%, 55%) |
|  | Unvaccinated | 152,460 | 183 | 11,056,080 | — | — |
| **Ages 12–15 years** |  |  |  |  |  |  |
| Medically diagnosed | BNT162b2 | 138,492 | 4,256 | 30,119,067 | 0.55 (0.52, 0.58) | 45% (42%, 48%) |
|  | Unvaccinated | 138,492 | 4,341 | 18,919,212 | — | — |
| Hospitalization/ED-diagnosed | BNT162b2 | 138,492 | 199 | 30,450,710 | 0.33 (0.27, 0.41) | 67% (59%, 73%) |
|  | Unvaccinated | 138,492 | 351 | 19,378,583 | — | — |
| **Ages 16–17 years** |  |  |  |  |  |  |
| Medically diagnosed | BNT162b2 | 70,365 | 2,233 | 16,631,210 | 0.49 (0.46, 0.52) | 51% (48%, 54%) |
|  | Unvaccinated | 70,365 | 2,604 | 10,375,472 | — | — |
| Hospitalization/ED-diagnosed | BNT162b2 | 70,365 | 134 | 16,829,911 | 0.31 (0.24, 0.39) | 69% (61%, 76%) |
|  | Unvaccinated | 70,365 | 257 | 10,681,097 | — | — |

— indicates the reference group; CI = confidence interval; COVID‑19 = coronavirus disease 2019; ED = emergency department; HR = hazard ratio; sIPTW = stabilized inverse probability of treatment weighted; VE = vaccine effectiveness.

eTable 4.	Estimated Vaccine Effectiveness of a Complete Primary Series of BNT162b2, Corrected for Potentially Missing Vaccine Records

|  | Optum | | CVS Health | |
| --- | --- | --- | --- | --- |
| COVID-19 Outcome | Hypothesized Sensitivity of Vaccination Exposure Measurement | Corrected VE (95% CI) | Hypothesized Sensitivity of Vaccination Exposure Measurement^a^ | Corrected VE (95% CI) |
| Medically diagnosed | 100% (original, uncorrected) | 35% (31%, 39%) | 100% (original, uncorrected) | 39% (37%, 41%) |
|  | 83% | 39% (35%, 43%) | 89% | 41% (39%, 43%) |
|  | 71% | 45% (41%, 48%) | 69% | 49% (48%, 51%) |
| Hospital/ED-diagnosed | 100% (original, uncorrected) | 55% (41%, 65%) | 100% (original, uncorrected) | 62% (57%, 67%) |
|  | 83% | 61% (49%, 70%) | 89% | 65% (60%, 69%) |
|  | 71% | 68% (58%, 75%) | 69% | 73% (69%, 76%) |

CI = confidence interval; COVID‑19 = coronavirus disease 2019; ED = emergency department; IIS = immunization information system; VE = vaccine effectiveness.

^a^ Hypothesized vaccination exposure measurements sensitivity were estimated by comparing the observed state-level vaccination coverages estimates in the linked claims-IIS data sources to estimates of statewide receipt of at least one COVID-19 vaccine dose from the Centers for Disease Control and Prevention, state departments of health, and capture-recapture methods.

eTable 5.	Estimated Pediatric Vaccine Effectiveness of a Complete Primary Series of BNT162b2, First 14 Days of Follow-up, Negative Control Analysis

| COVID-19 Outcome | Vaccine Exposure Group | N | Number of Events | Person-Time (Days) | sIPTW HR (95% CI) | sIPTW VE (95% CI) |
| --- | --- | --- | --- | --- | --- | --- |
| **Optum** |  |  |  |  |  |  |
| Medically diagnosed | BNT162b2 | 92,338 | 263 | 1,275,410 | 0.85 (0.72, 1.02) | 15% (-1.7%, 28%) |
|  | Unvaccinated | 92,338 | 278 | 1,188,068 | — | — |
| Hospitalization/ED-diagnosed | BNT162b2 | 92,338 | < 11 | 1,276,957 | 0.46 (0.17, 1.26) | 54% (-26%, 83%) |
|  | Unvaccinated | 92,338 | 12 | 1,189,854 | — | — |
| **CVS Health** |  |  |  |  |  |  |
| Medically diagnosed | BNT162b2 | 361,317 | 828 | 5,027,773 | 0.75 (0.68, 0.83) | 25% (17%, 32%) |
|  | Unvaccinated | 361,317 | 974 | 4,601,077 | — | — |
| Hospitalization/ED-diagnosed | BNT162b2 | 361,317 | 43 | 5,032,434 | 0.64 (0.43, 0.95) | 36% (5%, 57%) |
|  | Unvaccinated | 361,317 | 61 | 4,606,708 | — | — |

— indicates the reference group; CI = confidence interval; COVID‑19 = coronavirus disease 2019; ED = emergency department; HR = hazard ratio; VE = vaccine effectiveness.

Note: Optum privacy rules require masking cell sizes < 11 individuals.

eTable 6.	Distribution of Follow-up Person-time by Age Subgroup in Each Variant Era Analysis

##### A. Follow-up Time for Medically Diagnosed COVID-19

| Age Group | Person-time (Days) | | | | | |
| --- | --- | --- | --- | --- | --- | --- |
|  | Optum | | | CVS Health | | |
|  | Pre-delta | Delta | Omicron | Pre-delta | Delta | Omicron |
| **BNT162b2** |  |  |  |  |  |  |
| Overall | 769,372 (100%) | 3,699,202 (100%) | 709,984 (100%) | 3,058,485 (100%) | 14,616,762 (100%) | 2,412,356 (100%) |
| 5-11 years | 0 (0%) | 901,696 (24%) | 555,117 (78%) | 0 (0%) | 4,230,331 (29%) | 1,934,714 (80%) |
| 12-15 years | 245,740 (32%) | 2,191,191 (59%) | 110,646 (16%) | 935,058 (31%) | 8,181,097 (56%) | 349,760 (14%) |
| 16-17 years | 523,632 (68%) | 606,315 (16%) | 44,221 (6%) | 2,123,427 (69%) | 2,205,334 (15%) | 127,882 (5%) |
| **Unvaccinated** |  |  |  |  |  |  |
| Overall | 614,292 (100%) | 3,179,877 (100%) | 809,064 (100%) | 2,336,820 (100%) | 11,694,986 (100%) | 2,403,331 (100%) |
| 5-11 years | 0 (0%) | 726,740 (23%) | 594,804 (74%) | 0 (0%) | 3,170,004 (27%) | 1,868,530 (78%) |
| 12-15 years | 203,137 (33%) | 1,882,309 (59%) | 148,528 (18%) | 759,766 (33%) | 6,596,854 (56%) | 381,401 (16%) |
| 16-17 years | 411,155 (67%) | 570,828 (18%) | 65,732 (8%) | 1,577,054 (67%) | 1,928,128 (16%) | 153,400 (6%) |

COVID‑19 = coronavirus disease 2019.

Note: pre-delta era, December 11, 2020–May 31, 2021; delta era, June 1, 2021–December 24, 2021; omicron era, December 25, 2021–end of data availability.

##### B. Follow-up Time for Hospital/Emergency Department-Diagnosed COVID-19

| Age Group | Person-time (Days) | | | | | |
| --- | --- | --- | --- | --- | --- | --- |
|  | Optum | | | CVS Health | | |
|  | Pre-delta | Delta | Omicron | Pre-delta | Delta | Omicron |
| **BNT162b2** |  |  |  |  |  |  |
| Overall | 770,390 (100%) | 3,713,632 (100%) | 717,802 (100%) | 3,061,916 (100%) | 14,665,396 (100%) | 2,435,520 (100%) |
| 5-11 years | 0 (0%) | 905,166 (24%) | 561,458 (78%) | 0 (0%) | 4,239,598 (29%) | 1,954,605 (80%) |
| 12-15 years | 245,824 (32%) | 2,199,665 (59%) | 111,279 (16%) | 935,276 (31%) | 8,210,262 (56%) | 352,165 (14%) |
| 16-17 years | 524,566 (68%) | 608,801 (16%) | 45,065 (6%) | 2,126,640 (69%) | 2,215,536 (15%) | 128,750 (5%) |
| **Unvaccinated** |  |  |  |  |  |  |
| Overall | 615,697 (100%) | 3,219,630 (100%) | 819,005 (100%) | 2,343,319 (100%) | 11,837,049 (100%) | 2,433,893 (100%) |
| 5-11 years | 0 (0%) | 729,776 (23%) | 602,159 (74%) | 0 (0%) | 3,181,710 (27%) | 1,893,113 (78%) |
| 12-15 years | 203,261 (33%) | 1,911,495 (59%) | 150,313 (18%) | 760,047 (32%) | 6,693,946 (57%) | 385,957 (16%) |
| 16-17 years | 412,436 (67%) | 578,359 (18%) | 66,533 (8%) | 1,583,272 (68%) | 1,961,393 (17%) | 154,823 (6%) |

COVID‑19 = coronavirus disease 2019.

Note: pre-delta era, December 11, 2020–May 31, 2021; delta era, June 1, 2021–December 24, 2021; omicron era, December 25, 2021–end of data availability.

eTable 7.	Estimated Pediatric Vaccine Effectiveness of Receiving a Complete Primary Series of BNT162b2, by Era of Predominant Circulating Variants

##### A. Optum

| **COVID‑19 Outcome** | **Vaccine Exposure Group** | **N** | **Number of Events** | **Person-time (days)** | **sIPTW HR (95% CI)** | **sIPTW VE (95% CI)** |
| --- | --- | --- | --- | --- | --- | --- |
| **Pre-delta variant era** |  |  |  |  |  |  |
| Medically diagnosed | BNT162b2 | 31,050 | 36 | 769,372 | 0.46 (0.30, 0.71) | 54% (29%, 70%) |
|  | Unvaccinated | 31,050 | 66 | 614,292 | — | — |
| Hospital/ED–diagnosed | BNT162b2 | 31,050 | < 11 | 770,390 | 0.24 (0.04, 1.68) | 76% (-68%, 96%) |
|  | Unvaccinated | 31,050 | < 11 | 615,697 | — | — |
| **Delta variant era** |  |  |  |  |  |  |
| Medically diagnosed | BNT162b2 | 51,892 | 446 | 3,699,202 | 0.41 (0.37, 0.47) | 59% (53%, 63%) |
|  | Unvaccinated | 51,892 | 904 | 3,179,877 | — | — |
| Hospital/ED–diagnosed | BNT162b2 | 51,892 | 11 | 3,713,632 | 0.19 (0.10, 0.38) | 81% (62%, 90%) |
|  | Unvaccinated | 51,892 | 48 | 3,219,630 | — | — |
| **Omicron variant era** |  |  |  |  |  |  |
| Medically diagnosed | BNT162b2 | 9,396 | 130 | 709,984 | 0.92 (0.72, 1.17) | 8.2% (-17%, 28%) |
|  | Unvaccinated | 9,396 | 145 | 809,064 | — | — |
| Hospital/ED–diagnosed | BNT162b2 | 9,396 | < 11 | 717,802 | 0.40 (0.10, 1.54) | 60% (-54%, 90%) |
|  | Unvaccinated | 9,396 | < 11 | 819,005 | — | — |

— indicates the reference group; CI = confidence interval; COVID‑19 = coronavirus disease 2019; ED = emergency department; HR = hazard ratio; sIPTW = stabilized inverse probability of treatment weighted; VE = vaccine effectiveness.

Note: Optum privacy rules require masking cell sizes < 11 individuals.

##### B. CVS Health

| **COVID‑19 Outcome** | **Vaccine Exposure Group** | **N** | **Number of Events** | **Person-time (days)** | **sIPTW HR (95% CI)** | **sIPTW VE (95% CI)** |
| --- | --- | --- | --- | --- | --- | --- |
| **Pre-delta variant era** |  |  |  |  |  |  |
| Medically diagnosed | BNT162b2 | 118,851 | 119 | 3,058,485 | 0.37 (0.29, 0.47) | 63% (53%, 71%) |
|  | Unvaccinated | 118,851 | 245 | 2,336,820 | — | — |
| Hospital/ED–diagnosed | BNT162b2 | 118,851 | 4 | 3,061,916 | 0.41 (0.12, 1.36) | 59% (-36%, 88%) |
|  | Unvaccinated | 118,851 | 8 | 2,343,319 | — | — |
| **Delta variant era** |  |  |  |  |  |  |
| Medically diagnosed | BNT162b2 | 203,051 | 1,318 | 14,616,762 | 0.38 (0.35, 0.40) | 62% (60%, 65%) |
|  | Unvaccinated | 203,051 | 2,733 | 11,694,986 | — | — |
| Hospital/ED–diagnosed | BNT162b2 | 203,051 | 60 | 14,665,396 | 0.23 (0.17, 0.31) | 77% (69%, 83%) |
|  | Unvaccinated | 203,051 | 212 | 11,837,049 | — | — |
| **Omicron variant era** |  |  |  |  |  |  |
| Medically diagnosed | BNT162b2 | 39,415 | 509 | 2,412,356 | 0.90 (0.79, 1.02) | 10% (-2%, 21%) |
|  | Unvaccinated | 39,415 | 534 | 2,403,331 | — | — |
| Hospital/ED–diagnosed | BNT162b2 | 39,415 | 31 | 2,435,520 | 0.96 (0.58, 1.59) | 4% (-59%, 42%) |
|  | Unvaccinated | 39,415 | 32 | 2,433,893 | — | — |

— indicates the reference group; CI = confidence interval; COVID‑19 = coronavirus disease 2019; ED = emergency department; HR = hazard ratio; sIPTW = stabilized inverse probability of treatment weighted; VE = vaccine effectiveness.

eFigure 1.	Study Design and Variable Assessment Periods

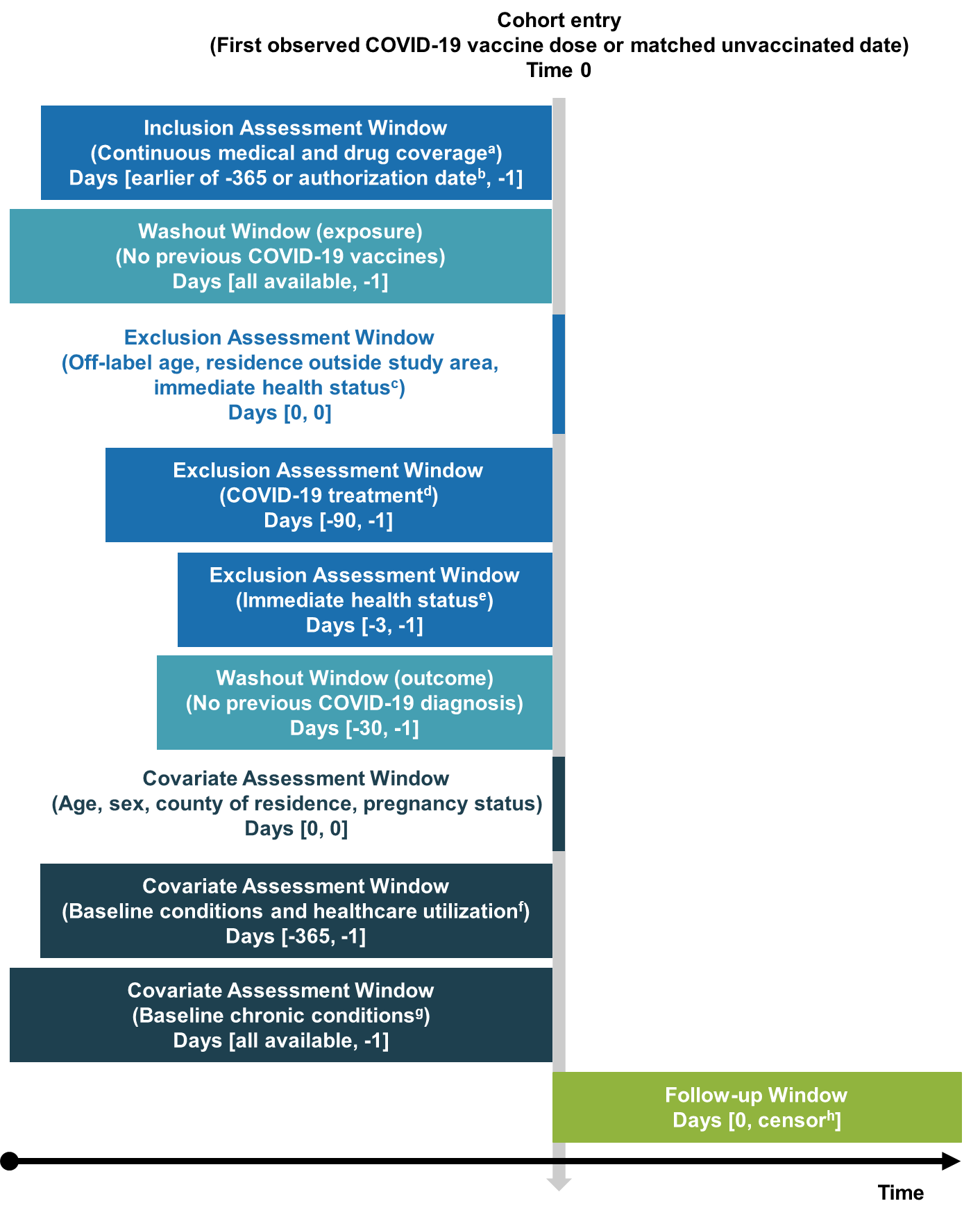

COVID-19 = coronavirus disease 2019; ED = emergency department.

^a^ Gaps in medical and pharmacy coverage < 32 days permitted.

^b^ December 11, 2020, ages 16–17 years; May 10, 2021, ages 12–17 years, October 29, 2021, ages 5–17 years.

^c^ Hospitalization or long-term care residence at Time 0.

^d^ COVID-19 monoclonal antibodies or convalescent plasma.

^e^ Diagnoses of general acute symptoms (fever, nausea/vomiting, rash) and health care utilization (hospitalization, ED visit) serving as an indicator of health status at the time of vaccination.

^f^ Number of hospitalizations, number of ED visits, skilled nursing facility stay, influenza vaccination, pneumococcal vaccination, encounter for cancer screening, eye examination, colonoscopy, bone mineral density test, well-check/well-child preventive health care visit, arthritis, lipid abnormality, ambulance use/life support service, weakness, pregnancy completion before Time 0.

^g^ Autoimmune disorders, cancer, chronic kidney disease or renal disease, chronic liver disease, chronic lung diseases (e.g., asthma, chronic obstructive pulmonary disease, cystic fibrosis, pulmonary embolism), dementia or other neurological conditions, diabetes mellitus type 1 or 2, Down syndrome, heart conditions (e.g., heart failure, coronary artery disease, arrhythmias), hypertension, immunocompromised state, mental health conditions, obese or severely obese, sickle cell disease or thalassemia, stroke or cerebrovascular disease, tuberculosis, COVID-19 laboratory test performed (binary indicator of any test performed or none), COVID-19 diagnoses.

^h^ End of study period, end of continuous health plan enrollment, relocation out of study area, deviation from the categorized vaccine exposure status (complete primary series or being unvaccinated).

eFigure 2.	Characteristics of Primary Series Completion

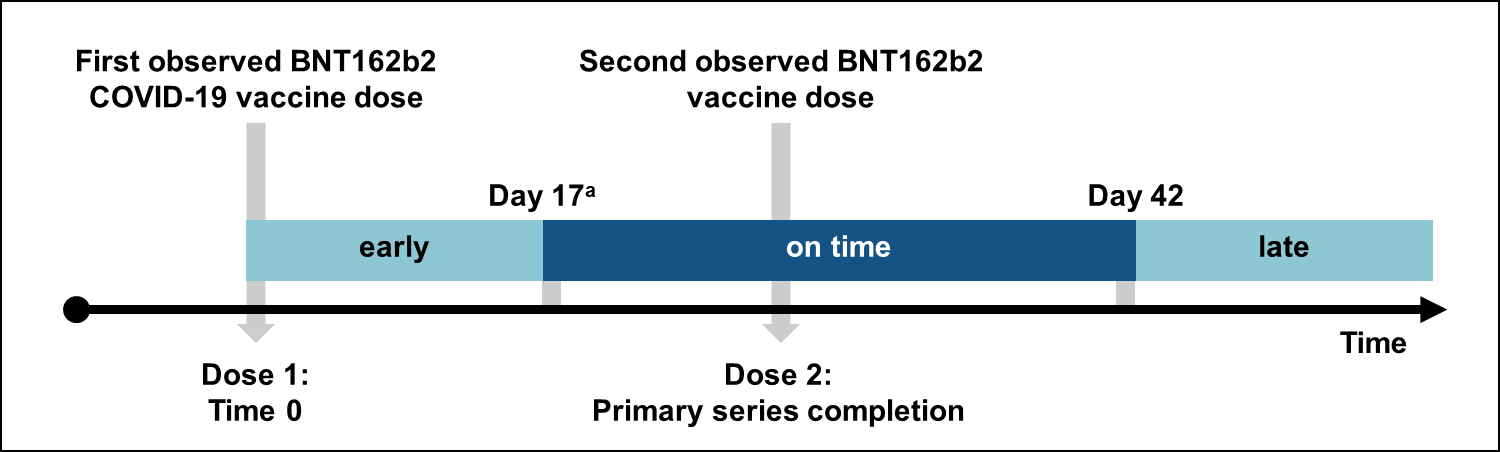

COVID-19 = coronavirus disease 2019.

^a^ 4 days before the recommended interval to received dose 2.

eFigure 3.	Attrition of the Analytic Cohort by Application of Eligibility Criteria.

##### A. Optum

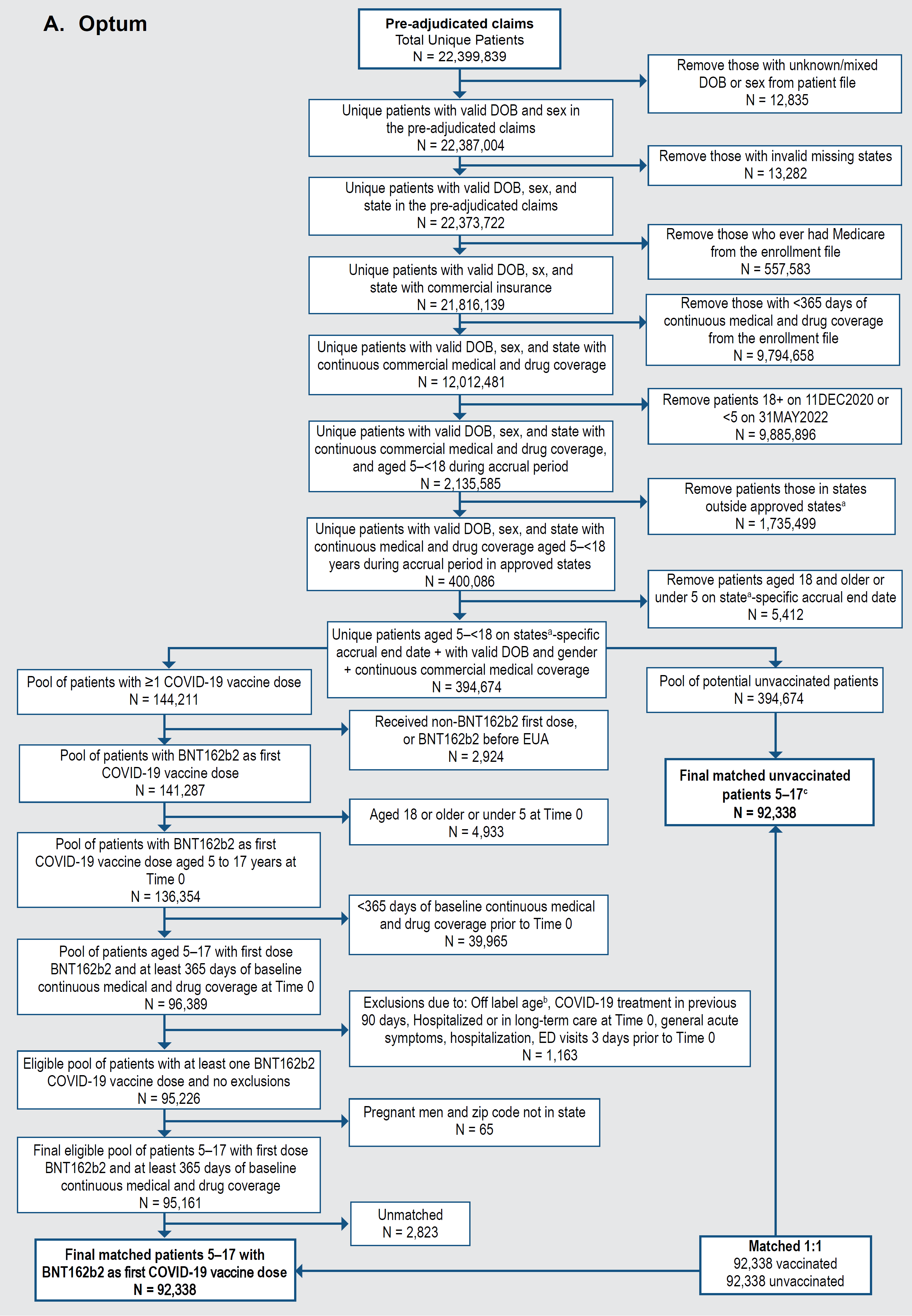

COVID-19 = coronavirus disease 2019; DOB = date of birth; IIS = immunization information system.

^a^ Ten IIS jurisdictions were included in the analysis, most of which were defined at the state level.

^b^ Children were removed if they received a COVID-19 vaccine when it was not authorized for their age group.

^c^ Children in the unvaccinated group may also be included in the vaccinated group with a different Time 0.

##### B. CVS Health

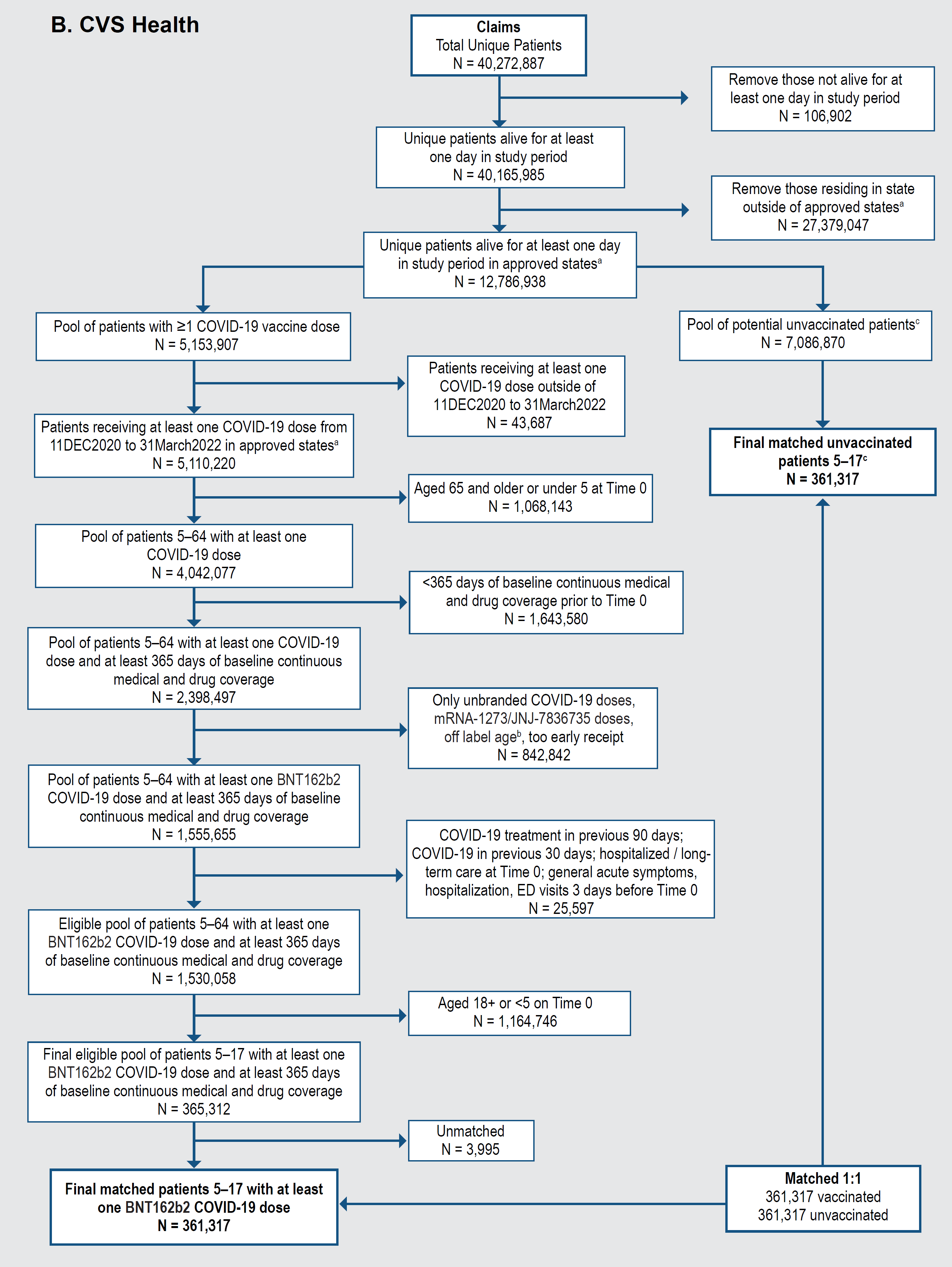

COVID-19 = coronavirus disease 2019; ED = emergency department.

^1^ Data from 11 jurisdictions from 9 states were used.

^2^ Patients were removed if they received a COVID-19 vaccine when it was not authorized for their age group.

^3^ Patients in the unvaccinated group may also join the vaccinated group.

eFigure 4.	Propensity Score Distributions in Matched BNT162b2 COVID-19 Vaccine Exposure Groups Among Children Aged 5–17 Years

##### A. Optum

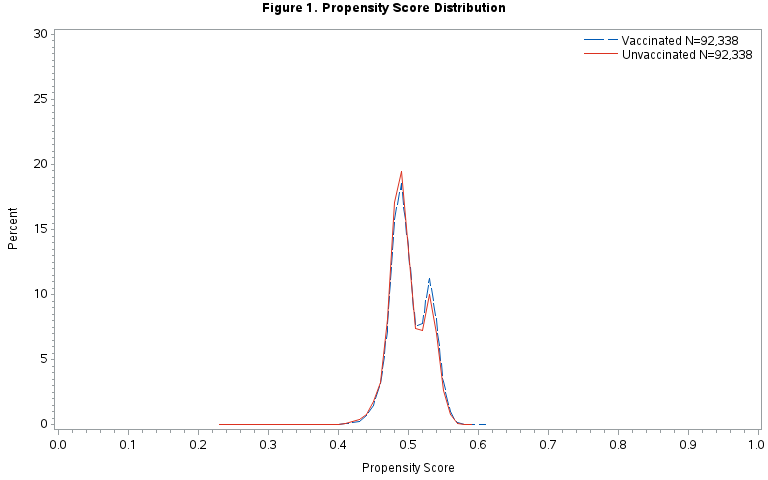

Note: propensity score variables included the following (in binary form except where indicated otherwise): age at Time 0 (linear); sex (categorical, reference = male); state (categorical); pregnant at Time 0; hospitalizations (categorical, reference = 0); emergency department visits (categorical, reference = 0); skilled nursing facility stay; influenza vaccination; pneumococcal vaccination; encounter for cancer screening; eye examination; colonoscopy; bone mineral density test; well-check/well-child preventive healthcare visit; arthritis; lipid abnormality; ambulance use or life support services; weakness; autoimmune disorders; cancer; chronic kidney disease or renal disease; chronic liver disease; chronic lung diseases (e.g., asthma, COPD, cystic fibrosis, pulmonary embolism); dementia or other neurological conditions; diabetes mellitus, type 1 or 2; down syndrome; heart conditions (e.g., heart failure, coronary artery disease, arrhythmias); hypertension; immunocompromised state; mental health conditions; obese or severely obese; sickle cell disease or thalassemia; stroke or cerebrovascular disease; tuberculosis; ≥ 1 COVID-19 laboratory test performed; COVID-19 diagnosis in any setting; delta or omicron variant era; having ≥ 1 condition increasing risk of COVID-19; delta era * COVID-19 laboratory test during baseline (interaction term).

##### B. CVS Health

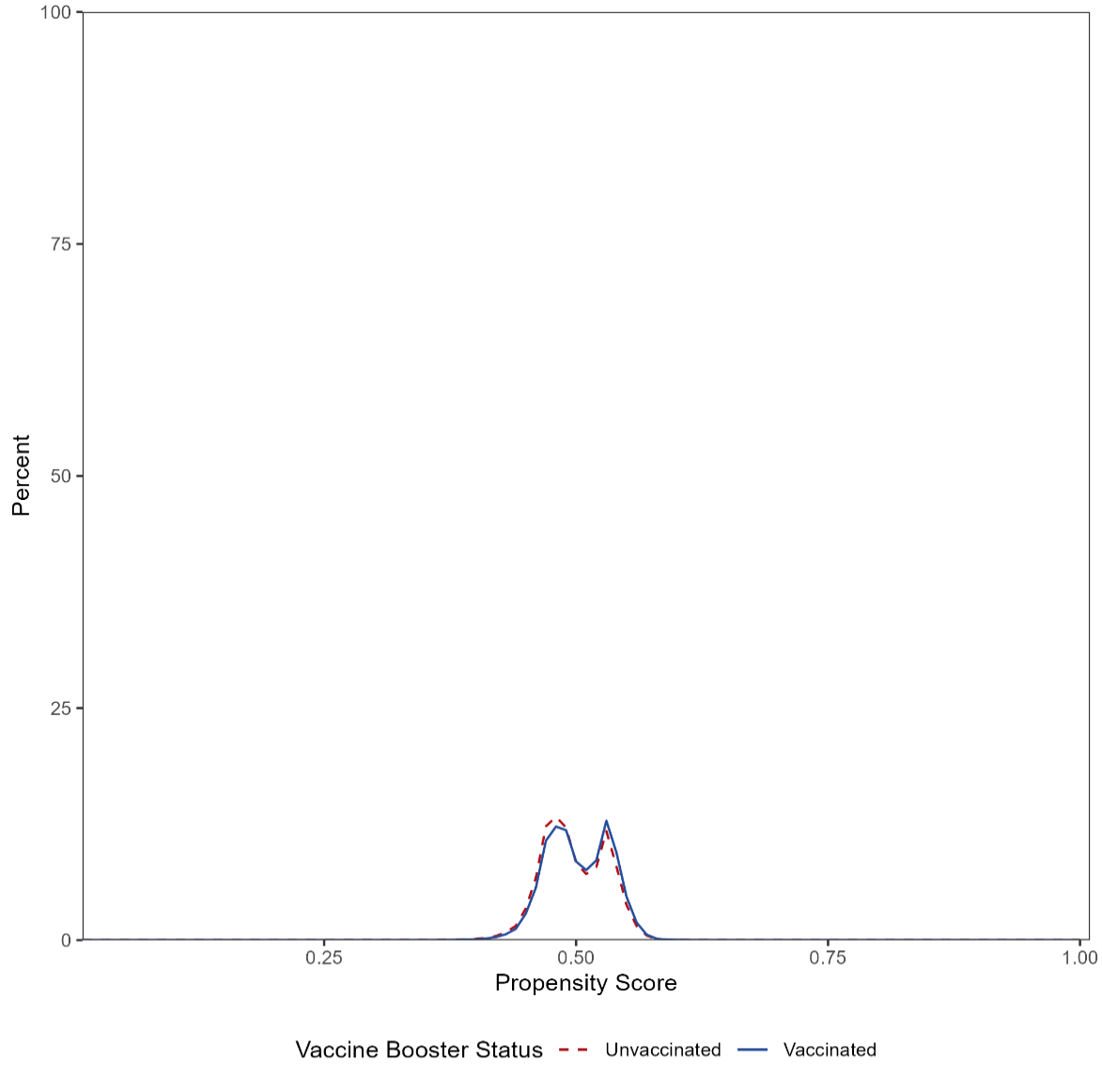

Note: propensity score variables included the following (in binary form except where indicated otherwise): age (categorical, reference = 5-11 years); ambulance use of life support services; arthritis; autoimmune disorders; bone mineral density test; cancer; encounter for cancer screening; chronic kidney disease or renal disease; chronic liver disease; colonoscopy; chronic lung diseases (e.g., asthma, COPD, cystic fibrosis, pulmonary embolism); pregnant at Time 0; COVID-19 diagnoses in any setting; hospitalization or emergency department-diagnosed COVID-19; ≥ 1 COVID-19 laboratory performed; COVID-19 diagnoses occurring outside of hospital or emergency department; COVID-19 vaccination index date in the delta/omicron era; diabetes mellitus, type 1 or 2; Down syndrome; emergency department visits (categorical, reference = 0); eye examination; influenza vaccination; heart conditions (e.g., heart failure, coronary artery disease, arrhythmias); ≥ 1 conditions which may qualify for priority groups for vaccination or booster dose eligibility; hypertension; hospitalizations (categorical, reference = 0); lipid abnormality; mental health conditions; dementia or other neurological conditions; obese or severely obese; pneumococcal vaccination; sex (categorical, reference = male); sickle cell disease or thalassemia; skilled nursing facility stay; state of residence (categorical); stroke or cerebrovascular disease; tuberculosis; immunocompromised state; pregnancy completion before Time 0; weakness; well-check/well-child preventive healthcare visit; COVID-19 vaccination after May 31, 2021 * ≥ 1 COVID-19 laboratory tests performed (interaction term).

eFigure 5.	Weighted Cumulative Incidence, First 14 Days After Time 0, Negative Control Outcome

##### A. Optum, Medically Diagnosed COVID-19

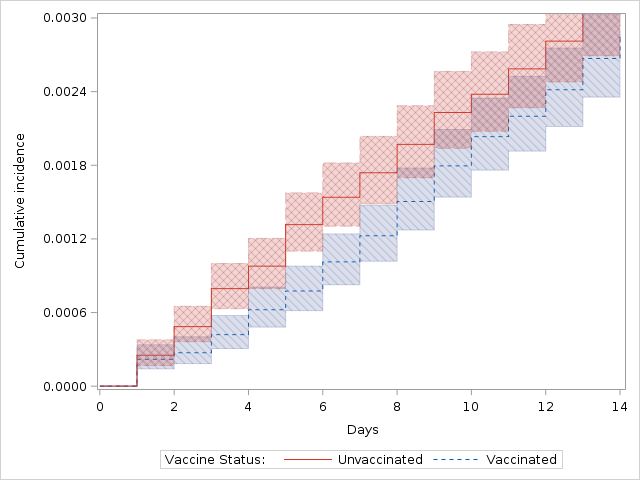

##### B. Optum, Hospital/ED–diagnosed COVID-19

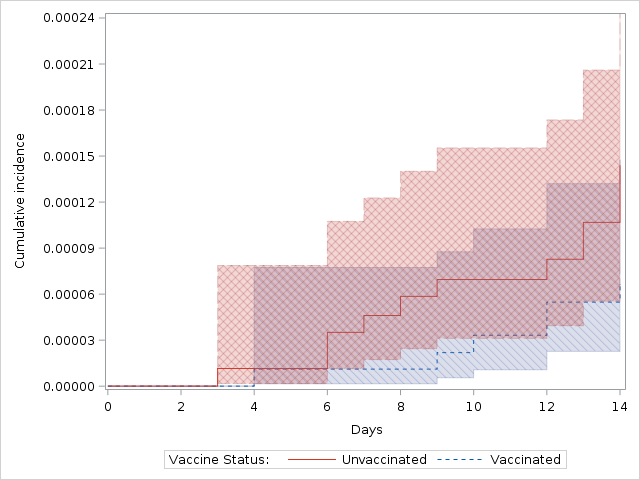

##### C. CVS Health, Medically Diagnosed COVID-19

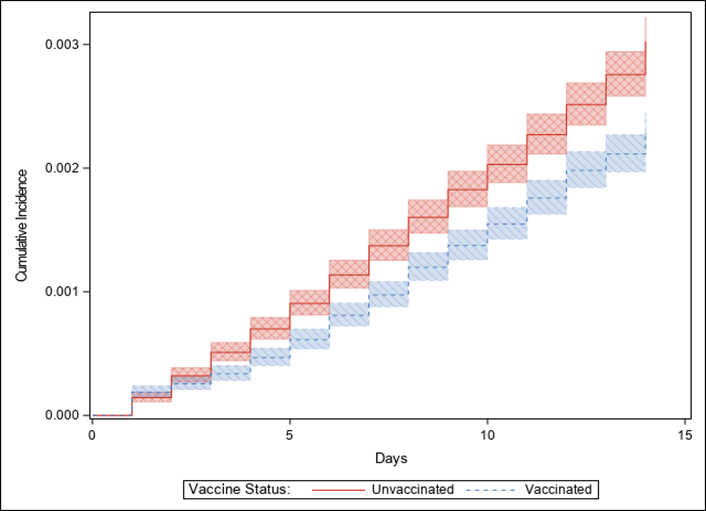

##### D. CVS Health, Hospital/ED–diagnosed COVID-19

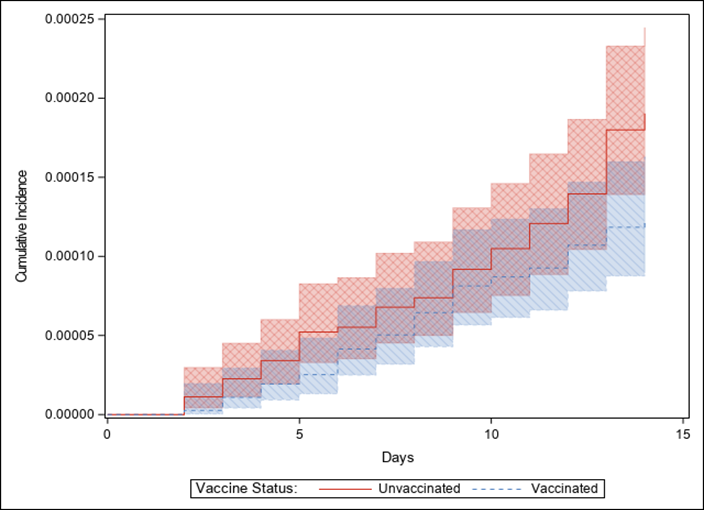

COVID-19 = coronavirus disease 2019; ED = emergency department.

eFigure 6.	Estimated Effectiveness of Receiving a Complete Primary Series of BNT162b2 in Children Aged 5 – 17 Years, Primary and Sensitivity Analyses

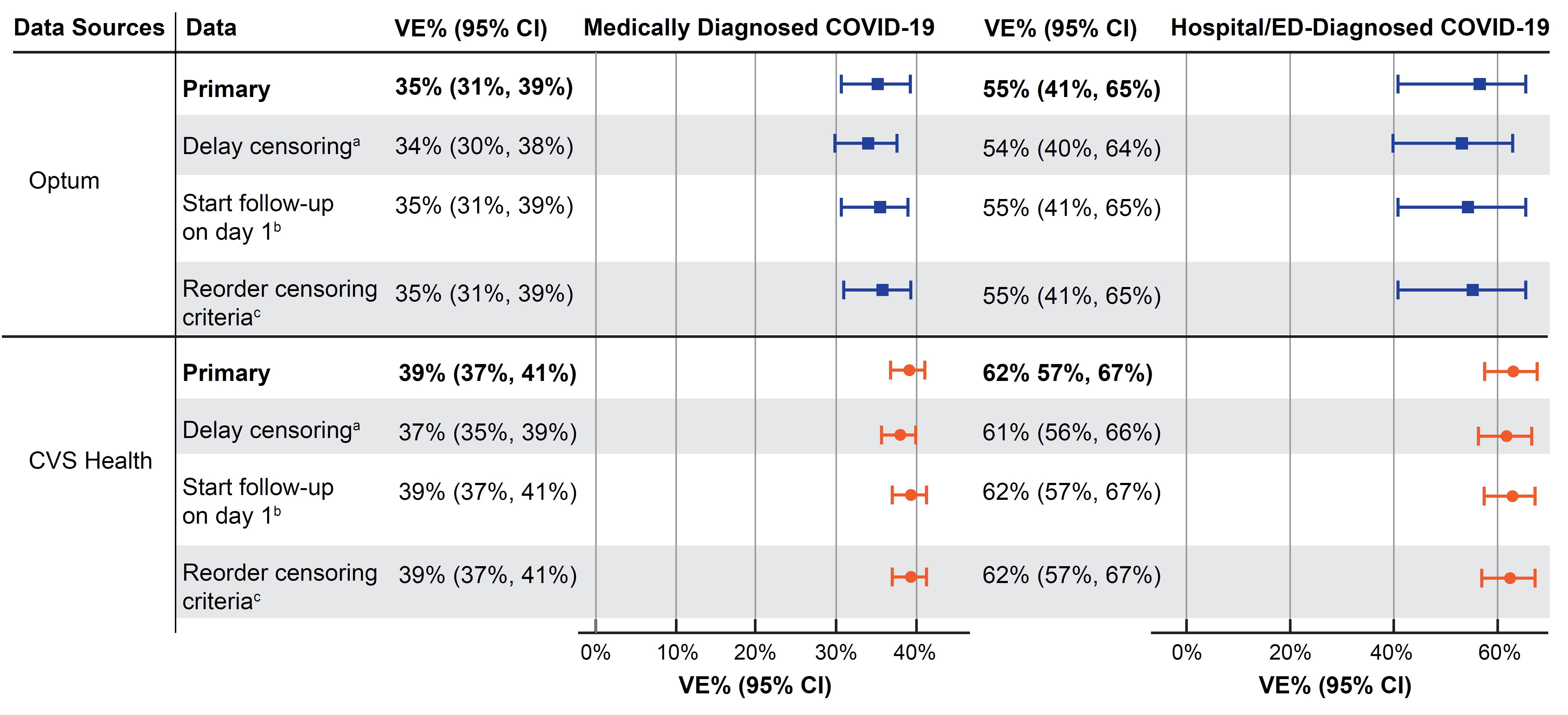

CI = confidence interval; COVID‑19 = coronavirus disease 2019; ED = emergency department; VE = vaccine effectiveness.

^a^ Censoring occurred 7 days after receipt of a censoring vaccine dose (i.e., in the vaccinated group: dose 2 received early, received dose 2 of a different brand; received dose 3; for unvaccinated, receipt of any dose) instead of censoring on the day of the vaccine dose.

^b^ Time 0 was removed from follow-up.

^c^ Censoring criteria were reordered so censoring for receipt of a censoring dose (i.e., in the vaccinated group: dose 2 received early, received dose 2 of a different brand; received dose 3; for unvaccinated, receipt of any dose) was applied before identifying outcomes or other censoring criteria on each day of follow-up (e.g., if a censoring vaccine dose and an outcome occurred on the same day, the child would be censored and not have the outcome counted).

eFigure 7.	Post Hoc Description of Daily COVID-19 Laboratory Testing by Vaccination Group During the Negative Control Period (Day 0–13), Optum

##### A. Overall

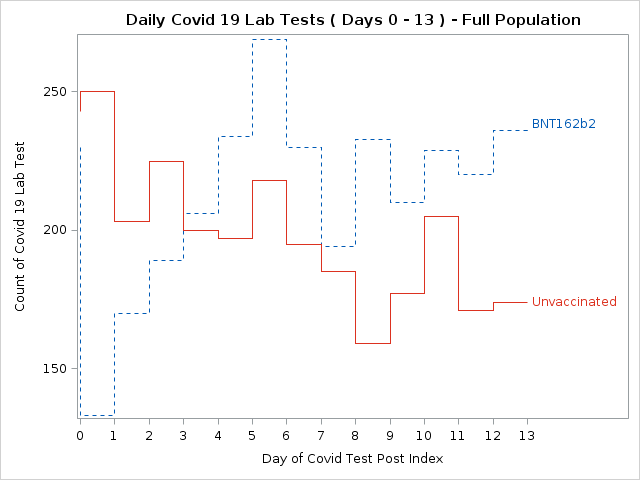

##### B. Among Those With a COVID-19 Diagnosis in Any Medical Setting in Days 0–13

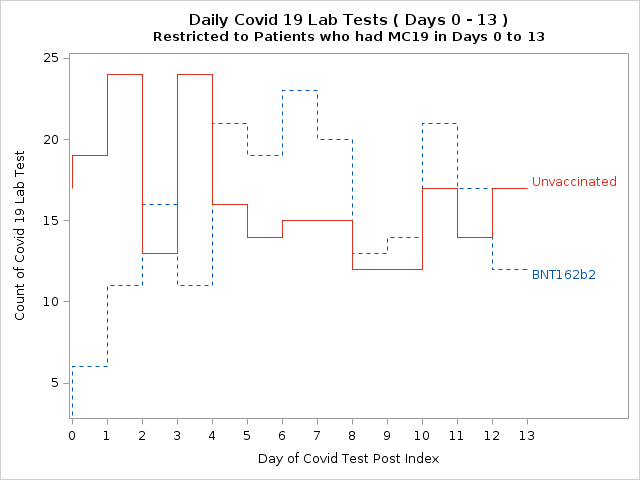

COVID-19 = coronavirus disease 2019.
